# Disparities in the pace of biological aging among midlife adults of the same chronological age have implications for future frailty risk and policy

**DOI:** 10.1101/2021.03.09.21252473

**Authors:** Maxwell L. Elliott, Avshalom Caspi, Renate M. Houts, Antony Ambler, Jonathan M. Broadbent, Robert J. Hancox, HonaLee Harrington, Sean Hogan, Ross Keenan, Annchen Knodt, Joan H. Leung, Tracy R. Melzer, Suzanne C. Purdy, Sandhya Ramrakha, Leah S. Richmond-Rakerd, Antoinette Righarts, Karen Sugden, W. Murray Thomson, Peter R. Thorne, Benjamin S. Williams, Graham Wilson, Ahmad R. Hariri, Richie Poulton, Terrie E. Moffitt

## Abstract

All humans age, but some age faster than others. Variation in biological aging can be measured in midlife, but the implications of this variation are poorly understood. We tested associations between biological aging and indicators of future frailty risk in the Dunedin cohort of 1037 infants born the same year and followed to age 45. Participants’ Pace of Aging was quantified by tracking declining function in 19 biomarkers indexing the cardiovascular, metabolic, renal, hepatic, immune, dental, and pulmonary systems across ages 26, 32, 38, and 45 years, in 2019. Participants with faster Pace of Aging had more cognitive difficulties, signs of advanced brain aging, diminished sensory-motor functions, older appearance, and more pessimistic perceptions of aging. People who are aging more rapidly than same-age peers in midlife may prematurely need supports to sustain independence that are usually reserved for older adults. Chronological age does not adequately identify need for such supports.

## INTRODUCTION

As we age, the risk that we will experience chronic diseases (e.g., heart disease, diabetes, cancer) and declining capacities (e.g., reduced strength, impaired hearing, worse memory) increases.^1^ To help mitigate personal and societal costs associated with aging, population-level policies typically specify eligibility on the basis of chronological age. These include retirement age, pensions, social security, and healthcare subsidies, all intended to support independence. However, while many individuals continue to live independently and flourish into their nineties, others experience organ failure, dementia, and mortality before their sixties, the age when entitlement to many of the aforementioned age-based supports begins.^2^ Thus, chronological age is, at best, an imperfect basis for aging policy.

All individuals age chronologically at the same rate, but there is marked variation in their rate of biological aging; this may help explain why some adults experience age-related decline faster than others.^3,4^ Biological aging can be defined as decline that (a) simultaneously involves multiple organ systems and (b) is gradual and progressive.^5^ Across the lifespan, the consequences of individual differences in genetic endowment, cellular biology, and life experiences accumulate, driving the divergence of biological age from chronological age for some people.^6–9^ Among older adults of the same chronological age, those with accelerated biological aging are more likely to develop heart disease, diabetes, and cancer and have a higher rate of cognitive decline, disability, and mortality.^10–16^

Current disease-management strategies usually treat and manage each age-related chronic disease independently.^7^ In contrast, the geroscience hypothesis proposes that many age-related chronic diseases could be prevented by slowing biological aging itself.^7,17^ The geroscience hypothesis states that biological aging drives cellular-level deterioration across all organ systems, thereby causing the exponential rise in multi-morbidity across the second half of the lifespan.^6^ The implication is that by slowing biological aging directly, instead of managing each disease separately, risk for all chronic age-related diseases could be simultaneously ameliorated.^5^ Early trials suggest that this goal may be attainable.^18,19^ To achieve maximal prevention of age-related diseases, interventions to slow biological aging will need to target individuals by midlife before decades of subclinical organ decline have accumulated.^6,20^ However, little is known about how to identify adults in midlife who are aging fast and who are most likely to benefit from geroscience-informed interventions, and for this reason, we studied biological aging in midlife.

We measured biological aging in a population-representative 1972–1973 birth cohort of 1,037 individuals followed from birth to age 45 years in 2019 with 94% retention: the Dunedin Study.^21^ Over a 20-year period—at ages 26, 32, 38, and 45—we repeatedly collected 19 biomarkers to assess changes in the function of cardiovascular, metabolic, renal, hepatic, immune, dental, and pulmonary systems, and quantified age-related decline shared among these systems (**Figure 1**). We call this index of biological aging in the Dunedin Study the “Pace of Aging”. We tested the hypothesis that individual differences in the Pace of Aging from ages 26 to 45 would be associated, at age 45, with established risk factors for future frailty, morbidity and early mortality across four domains (**Figure 1**).^22^ First, we tested whether individuals with a faster Pace of Aging had early signs of brain aging that have been linked to cognitive decline and dementia in older adults. Second, we tested whether individuals with a faster Pace of Aging had more cognitive difficulties and decline. Third, we tested whether those with a faster Pace of Aging already displayed signs of diminished sensory-motor functional capacities that are linked to loss of independence, falls, and mortality in studies of older adults. Fourth, we tested whether individuals with an accelerated Pace of Aging look older than their same-aged peers, whether they self-report pessimism about aging, and whether informants have noticed age-related difficulties in Study members.

**Figure 1.**
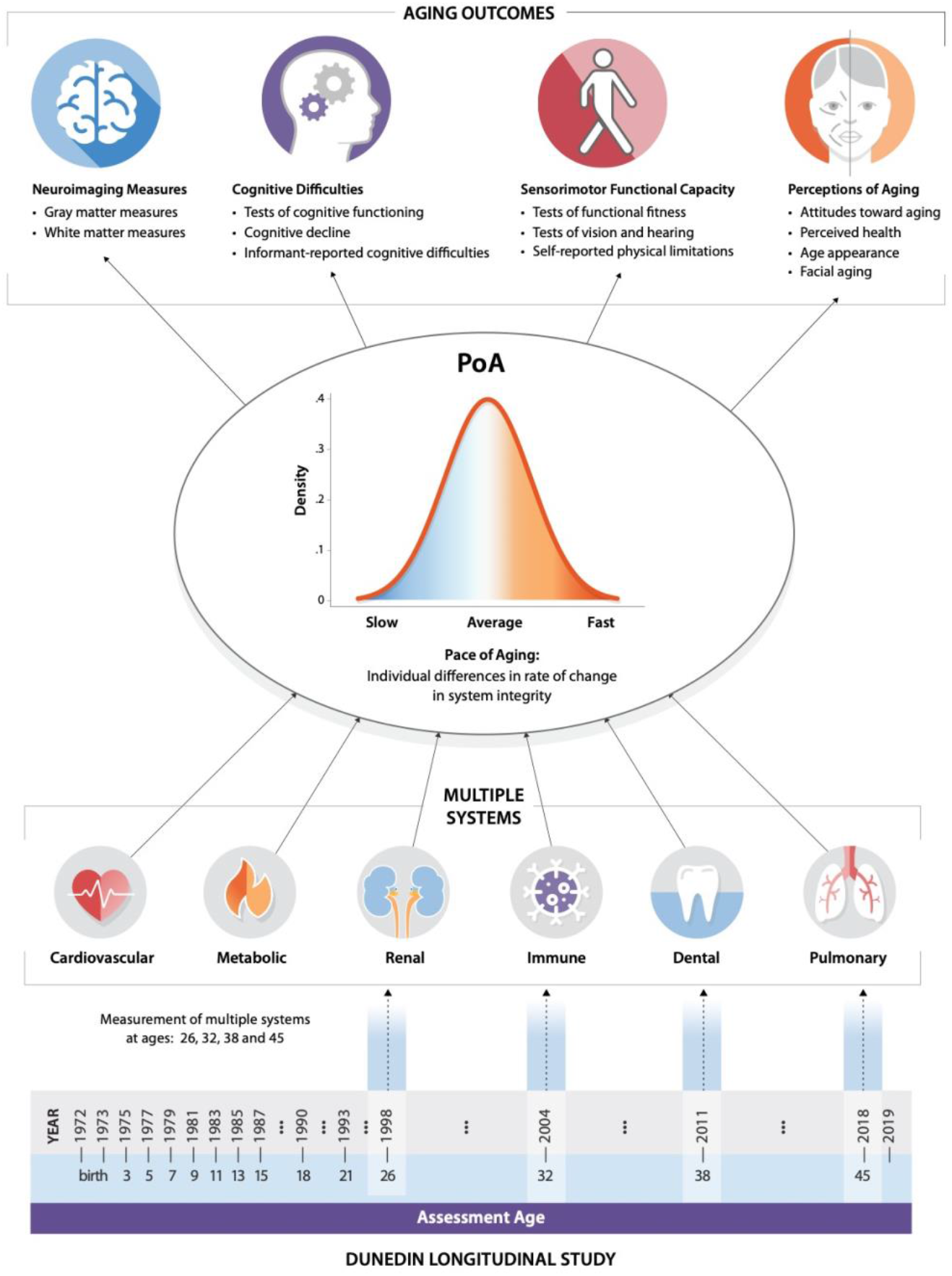
Study design. We studied the Pace of Aging in the Dunedin birth cohort. The timeline on the bottom of the figure visualizes the design of the Dunedin Longitudinal Study. The years of each phase of data collection and the corresponding ages are listed. The Pace of Aging was derived from measuring longitudinal changes in 19 biomarkers at 4 timepoints between ages 26 and 45 years. These biomarkers indexed functioning across multiple organ systems (each visualized under the heading “multiple systems”). We combined rates of changes across these biomarkers to produce a single measure termed the Pace of Aging (PoA). We then investigated associations between the Pace of Aging and aging outcomes across 4 domains at age 45: Neuroimaging measures, cognitive difficulties, sensorimotor functional capacity and perceptions of aging.

## RESULTS

### Quantifying two decades of biological aging in midlife

In a 2015 article, we quantified the Pace of Aging across 12 years among Dunedin Study members, from age 26 to age 38.^22^ Here, we extended these measurements to age 45, quantifying 20 years of biological aging across the first half of the lifespan (see **Supplemental Table S1** for details on each biomarker). From these biomarkers, the Pace of Aging was quantified in three steps.

First, we measured longitudinal changes in 19 biomarkers at ages 26, 32, 38 and 45 assessing cardiovascular, metabolic, renal, hepatic, immune, dental, and pulmonary systems, totaling 69,715 data points (cohort participants x biomarkers x assessment phases) (**Figure 1**). All biomarkers at each age were standardized based on their original distribution at age 26 (i.e. set to mean of 0 and a standard deviation of 1) and coded so that higher values represented “older/less healthy” levels (i.e., scores were reversed for cardiovascular fitness, lung function, creatinine clearance, and high density lipoprotein cholesterol for which values are expected to decline with increasing chronological age). In our cohort of midlife adults, biomarkers showed a pattern of age-dependent decline in the functioning of multiple organ systems over the 20-year follow-up period.

Second, linear mixed-effects modelling was used to quantify each study member’s personal rate of change across each of the 19 biomarkers. The 19 models took the form *B*_*it*_ = γ_0_ +γ_1_Age_*it*_ +μ_0i_ +μ_1i_Age_*it*_ + ϵ_*it*_, where *B*_*it*_ is a biomarker measured for individual *i* at time *t*, γ_0_ and γ_1_ are the fixed intercept and slope estimated for the cohort, and μ_0*i*_ and μ_1*i*_ are the random intercepts and slopes estimated for each individual *i*. Biomarker slopes indicated a tendency to decline with age (**Figure 2A**). Of the 171 unique correlations among biomarker slopes, 124 (73%) had a positive sign indicating coordinated change with age. Correlations between biomarker slopes averaged r = 0.1 ranging from r = −0.2 to r = 0.7 across the 19 biomarkers (**Supplemental Table S2**).

**Figure 2.**
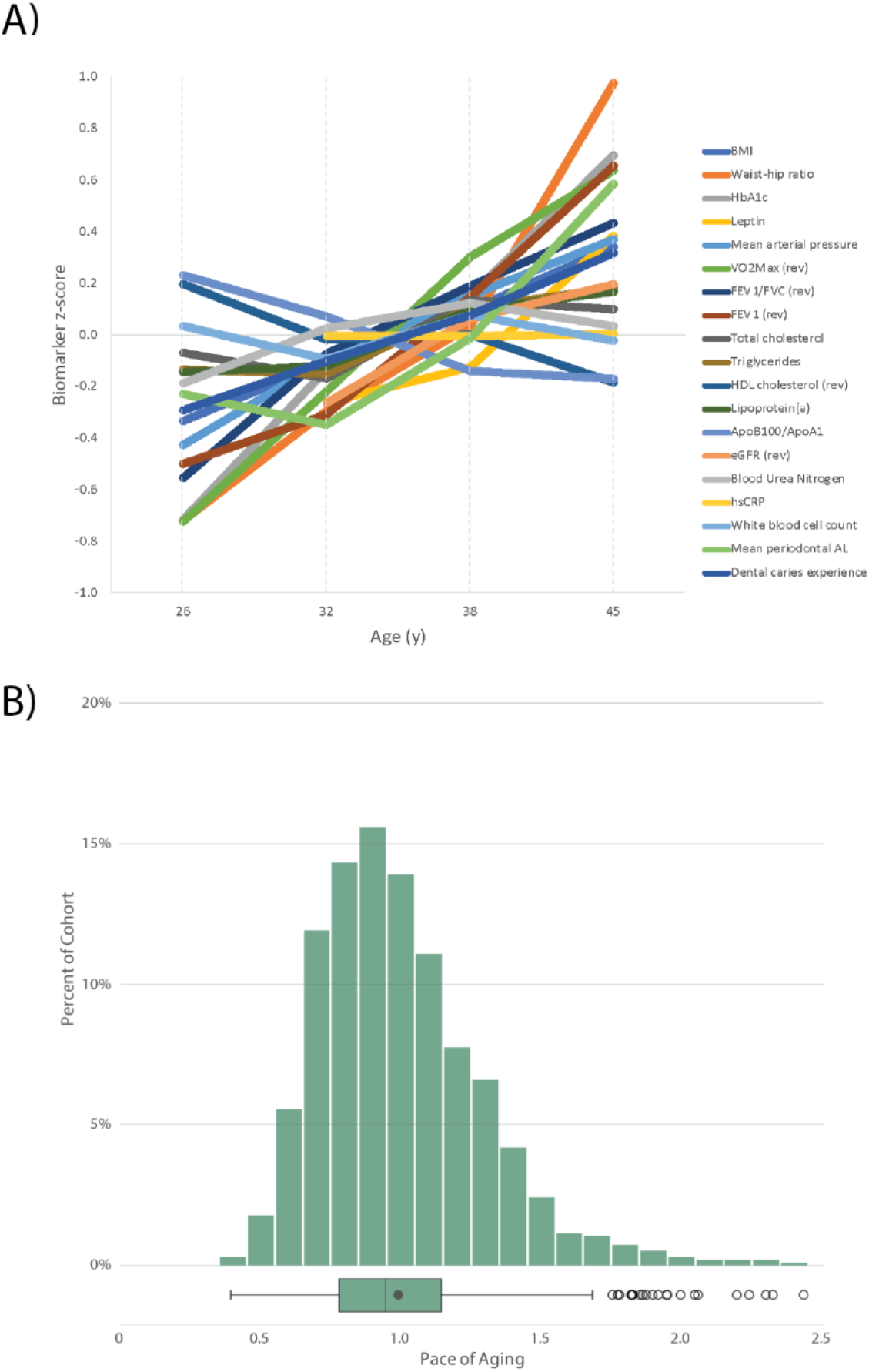
Biological aging across two decades from age 26 to age 45. A) For visualization, biomarker values were standardized to have M = 0 and SD = 1 across the two decades of follow-up (z-scores). Z-scores were coded so that higher values corresponded to older levels of the biomarkers. B) Pace of Aging is denominated in years of physiological change per chronological year. A Pace of Aging of one indicates a cohort member who experienced one year of physiological change per chronological year (the cohort average). A Pace of Aging of two indicates a cohort member aging at a rate of two years of physiological change per chronological year (i.e., twice as fast as the cohort average).

Third, we combined information from the 19 slopes to calculate each Study member’s personal Pace of Aging. In line with the geroscience hypothesis, which states that aging represents correlated gradual decline across organ systems, we calculated each Study member’s Pace of Aging as the sum of age-dependent annual changes across all biomarkers: 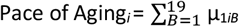. The resulting Pace of Aging was then scaled to a mean of 1, so that it could be interpreted with reference to an average rate of 1 year of biological aging per year of chronological aging. Study members showed wide variation in their Pace of Aging (Mean=1 biological year per chronological year, SD=0.29). Over the two decades that we measured biological aging, the Study member with the slowest Pace of Aging aged by just 0.40 biological years per chronological year, while the Study member with the fastest Pace of Aging accrued 2.44 biological years per chronological year **(Figure 2B)**.

### Accelerated biological aging and the aging brain

Deterioration of the brain (e.g., in Alzheimer’s disease and related dementias) is a major contributor to morbidity and loss of independence in older adults.^23,24^ Brain imaging can detect subtle signs of brain aging decades before the onset of age-related disease.^25,26^ Several magnetic resonance imaging (MRI) measures have been associated with a higher risk for cognitive decline and neurodegenerative disease in older adults including: thinner cortex, smaller surface area, smaller hippocampal volume, larger volume of white matter hyperintensities, lower fractional anisotropy, and older brain age.^27–29^ Here we found that an accelerated Pace of Aging in the first half of the lifespan was associated with most of these risk factors derived from high-resolution structural MRI scans at age 45. **Table 1** reports effect sizes, significance tests and covariate-adjusted analyses for all brain measures.

**Table 1.** Associations between the Pace of Aging, neuroimaging and cognitive measures. On the left side of this table are associations from the main text from regression models that were adjusted for sex. In the middle are sensitivity analyses in which the models also adjusted for BMI and smoking. On the right are results from models adjusted for sex in which all Study members who had a diagnosis of cancer, diabetes or heart attack were excluded (N=58. RAVL = Rey Auditory Verbal Learning. All statistically significant (p < .05) sex-adjusted associations remain statistically significant after false-discovery rate correction for the 38 tests presented in Table 1 and Table 2.

| Signs of brain aging and cognitive difficulties |  |  |  |  |  |  |  |  |  |
| --- | --- | --- | --- | --- | --- | --- | --- | --- | --- |
|  | Adjusted for sex |  |  | Adjusted for sex, BMI and smoking |  |  | Without cancer, diabetes or heart attack |  |  |
| | N | $\beta$ (95% CI) | P | N | $\beta$ (95% CI) | P | N | $\beta$ (95% CI) | P |
| **Signs of brain aging** |  |  |  |  |  |  |  |  |  |
| Cortical thickness | 860 | -0.14 (-0.21 to -0.08) | <.001 | 858 | -0.15 (-0.24 to -0.05) | .002 | 806 | -0.14 (-0.22 to -0.07) | < 0.001 |
| Surface area | 860 | -0.08 (-0.14 to -0.03) | .003 | 858 | -0.13 (-0.20 to -0.05) | .001 | 806 | 0.06 (-0.12 to -0.01) | 0.031 |
| Hippocampal volume | 860 | -0.10 (-0.16 to -0.04) | .001 | 858 | -0.14 (-0.22 to -0.06) | .001 | 806 | -0.08 (-0.15 to -0.02) | 0.009 |
| Log WMH volume | 851 | 0.18 (0.11 to 0.24) | <.001 | 848 | 0.17 (0.07 to 0.26) | <.001 | 797 | 0.18 (0.12 to 0.26) | < 0.001 |
| Fractional Anisotropy | 853 | -0.03 (-0.09 to 0.04) | .439 | 852 | -0.12 (-0.22 to -0.03) | .010 | 802 | -0.02 (-0.09 to 0.05) | 0.620 |
| BrainAGE | 868 | 0.20 (0.13 to 0.26) | <.001 | 865 | 0.20 (0.11 to 0.29) | <.001 | 813 | 0.18 (0.11 to 0.25) | < 0.001 |
| **Cognitive difficulties** |  |  |  |  |  |  |  |  |  |
| Full-scale IQ | 916 | -0.33 (-0.38 to -0.26) | <.001 | 910 | -0.32 (-0.41 to -0.24) | <.001 | 859 | -0.33 (-0.40 to -0.27) | < 0.001 |
| IQ decline (residualized change) | 904 | -0.16 (-0.22 to -0.09) | <.001 | 899 | -0.18 (-0.27 to -0.09) | <.001 | 847 | -0.17 (-0.25 to -0.11) | < 0.001 |
| Verbal comprehension index | 893 | -0.30 (-0.36 to -0.24) | <.001 | 889 | -0.35 (-0.44 to -0.26) | <.001 | 838 | -0.31 (-0.39 to -0.25) | < 0.001 |
| Perceptual reasoning index | 904 | -0.27 (-0.33 to -0.20) | <.001 | 900 | -0.27 (-0.36 to -0.18) | <.001 | 848 | -0.26 (-0.34 to -0.20) | < 0.001 |
| Working memory index | 900 | -0.22 (-0.28 to -0.15) | <.001 | 896 | -0.18 (-0.27 to -0.09) | <.001 | 844 | -0.22 (-0.30 to -0.16) | < 0.001 |
| Processing speed index | 904 | -0.23 (-0.29 to -0.16) | <.001 | 900 | -0.24 (-0.33 to -0.15) | <.001 | 848 | -0.22 (-0.29 to -0.16) | < 0.001 |
| RAVL learning memory | 905 | -0.29 (-0.34 to -0.22) | <.001 | 901 | -0.28 (-0.37 to -0.20) | <.001 | 849 | -0.21 (-0.28 to -0.15) | < 0.001 |
| RAVL recall | 901 | -0.19 (-0.25 to -0.13) | <.001 | 897 | -0.20 (-0.29 to -0.12) | <.001 | 845 | -0.30 (-0.37 to -0.25) | < 0.001 |
| Informant memory difficulties | 881 | 0.15 (0.08 to 0.21) | <.001 | 875 | 0.24 (0.15 to 0.33) | <.001 | 827 | 0.16 (0.10 to 0.24) | < 0.001 |
| Informant attention difficulties | 881 | 0.20 (0.14 to 0.26) | <.001 | 875 | 0.29 (0.20 to 0.38) | <.001 | 827 | 0.22 (0.16 to 0.29) | < 0.001 |

Study members with a faster Pace of Aging had thinner average cortical thickness (β = −0.14, p < 0.001; 95% CI: −0.21, −0.08) and smaller total surface area of the cortex (β = −0.08, p = 0.003; 95% CI: − 0.14, −0.03). Furthermore, regional investigation of cortical thickness revealed that associations between faster Pace of Aging and thinner cortex were widespread across the cortex (89.72% of parcels had negative effect sizes, 38.33% were statistically significant at p < .05, corrected for false discovery rate; **Supplemental Table S3**), with the largest associations in medial temporal and insular cortex (**Figure 3A**). Regional associations with surface area were also widespread across the cortex (96.11% of parcels had negative effect sizes, 22.50% were statistically significant p < .05, corrected for false discovery rate; **Supplemental Table S4**), with the largest associations in visual and lateral temporal cortex (**Figure 3B**). Those with a faster Pace of Aging also had smaller volumes of the hippocampus (β = −0.10, p = 0.001; 95% CI: −0.16, −0.04), a brain region central to both healthy memory function and age-related memory decline.^30^

**Figure 3.**
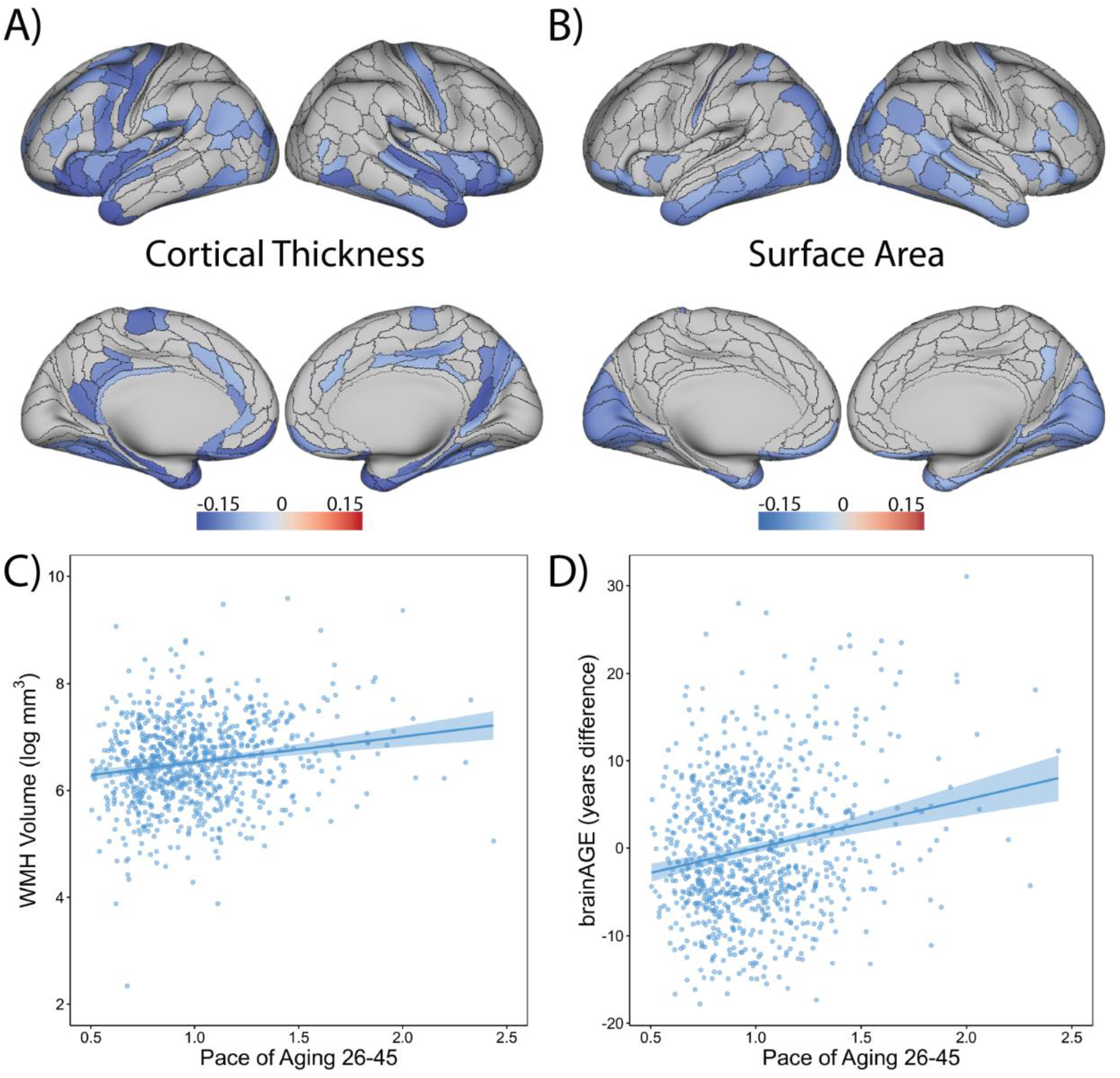
Study members who were aging faster showed signs of advanced brain aging relative to slower-aging peers. The overlays display cortical regions (in blue) whose (A) thickness or (B) surface area are significantly associated (p < 0.05, false discovery rate corrected) with Pace of Aging. The scatter plots show associations between Pace of Aging and (C) volume of white matter hyperintensities (WMH) as well as (D) brainAGE (a measure of the difference between each Study member’s chronological age and their brain age as estimated from a machine-learning algorithm that was trained to predict chronological age from gray- and white-matter measures in independent samples ranging in age from 19 to 82.^31^ Scatterplots include regression lines along with 95% confidence intervals.

Study members with a faster Pace of Aging had early signs of white matter deterioration, as indicated by a larger log-transformed volume of white matter hyperintensities (β = 0.18, p < 0.001; 95% CI: 0.11, 0.24; **Figure 3C**), but they did not have lower fractional anisotropy (β = −.03, p = 0.439; 95% CI: − 0.09, 0.04), a measure of white matter microstructural integrity. Sensitivity analyses revealed that associations between Pace of Aging and MRI signs of brain aging were not attributable to being overweight, to smoking history, or to already being diagnosed with an age-related disease (heart disease, diabetes, cancer) (**Table 1**).

We also studied a relatively new measure called “brain Age Gap Estimate” (brainAGE). BrainAGE is the difference between each study member’s chronological age and their brain age as estimated from a machine-learning algorithm that was trained to predict chronological age from gray-and white-matter measures in independent samples ranging in age from 19 to 82.^31^ Higher scores on brainAGE thus indicate a brain age that is older than chronological age. Dunedin Study members with a faster Pace of Aging tended to have brains that were typical of an older person as represented by higher brainAGE scores (β = 0.20, p < 0.001; 95% CI: 0.13, 0.26; **Figure 3D)**.

### Accelerated biological aging, cognitive difficulties and cognitive decline

Cognitive testing is widely used to assess risk for age-related neurological disease. Low cognitive functioning is a risk factor for Alzheimer’s disease and dementia, and cognitive decline is a hallmark feature of these age-related disorders.^32,33^ Dunedin Study members with a faster Pace of Aging displayed poorer cognitive functioning and more cognitive decline by age 45. **Table 1** reports effect sizes, significance tests and covariate-adjusted analyses for all cognitive measures.

Compared to peers with a slower Pace of Aging, those who were aging faster had lower intelligence quotient (IQ) scores (β = −0.33, p < .001; 95% CI: −0.38, −0.26). This difference in cognitive functioning reflected actual cognitive decline over the years: when we compared age-45 IQ test scores with baseline scores from the childhood version on the same IQ test, Study members with a faster Pace of Aging tended to show decline net of their baseline level (β = −0.16, p < 0.001; 95% CI: −0.22, −0.09). Furthermore, a faster Pace of Aging was broadly associated with poorer cognitive functioning across domains: Study members with a faster Pace of Aging had poorer verbal comprehension (β = −0.30, p < 0.001; 95% CI: −0.36, −0.24), perceptual reasoning (β = −0.27, p < 0.001; 95% CI: −0.33, −0.20), working memory (β = −0.22, p < 0.001; 95% CI: −0.28, −0.15), processing speed (β = −0.23, p < 0.001; 95% CI: −0.29, −0.16), worse memory learning performance (Rey Auditory Verbal Learning [RAVL] learning memory, β = −0.29, p < 0.001; 95% CI: −0.34, −0.22), and worse delayed memory recall (RAVL recall, β = −0.19, p < 0.001; 95% CI: −0.25, −0.13).

Cognitive difficulties were not only detectable on objective tests but also noticeable in everyday life. Informants, who were surveyed because they knew a Study member well, reported that Study members with a faster Pace of Aging experienced more memory difficulties (β = 0.15, p < 0.001; 95% CI: 0.08, 0.21) and attention problems (β = 0.20, p < 0.001; 95% CI: 0.14, 0.26); for example, they noted that faster-aging Study members were more likely to be “easily distracted” and “get sidetracked” as well as to “misplace wallet, keys or eyeglasses” and “forget to do errands, return calls or pay bills.” Sensitivity analyses indicated that the associations between Pace of Aging and cognitive difficulties were not attributable to being overweight, to smoking or to common age-related diseases (**Table 1**).

### Accelerated biological aging and diminished sensory-motor functional capacities

In gerontology, poor scores on tests of sensory-motor functioning (e.g., gait speed, grip strength, visual contrast sensitivity, hearing thresholds) are often used to identify frail individuals who are at high risk for falls, loss of independence, and mortality.^34–37^ Dunedin Study members who were aging faster showed multiple signs of sensory-motor difficulties. **Table 2** reports effect sizes, significance tests and covariate-adjusted analyses for all sensory-motor measures.

**Table 2.** Associations between the Pace of aging, measures of sensory-motor functional capacity and perceptions of aging. On the left side of this table are associations from the main text from regression models that were adjusted for sex. On the right side of the table are sensitivity analyses in which the models also adjusted for BMI and smoking. On the right are results from models adjusted for sex in which all Study members who had a diagnosis of cancer, diabetes or heart attack were excluded (N=58). All statistically significant (p < .05) associations in this table remain statistically significant after false-discovery rate correction for the 38 tests presented in Table 1 and Table 2.

| Sensory-motor functional capacity and perceptions of aging |  |  |  |  |  |  |  |  |  |
| --- | --- | --- | --- | --- | --- | --- | --- | --- | --- |
|  | Adjusted for sex |  |  | Adjusted for sex, BMI and smoking |  |  | Without cancer, diabetes or heart attack |  |  |
| | N | $\beta$ (95% CI) | P | N | $\beta$ (95% CI) | P | N | $\beta$ (95% CI) | P |
| **Sensory-motor capacity** |  |  |  |  |  |  |  |  |  |
| Gait speed | 903 | -0.33 (-0.39 to -0.27) | <.001 | 901 | -0.19 (-0.27 to -0.10) | <.001 | 848 | -0.33 (-0.40 to -0.27) | < 0.001 |
| One-legged balance | 909 | -0.36 (-0.42 to -0.30) | <.001 | 905 | -0.26 (-0.34 to -0.17) | <.001 | 853 | -0.35 (-0.42 to -0.29) | < 0.001 |
| Chair stands | 872 | -0.30 (-0.37 to -0.24) | <.001 | 871 | -0.25 (-0.34 to -0.16) | <.001 | 820 | -0.30 (-0.38 to -0.24) | < 0.001 |
| Step in place | 885 | -0.28 (-0.34 to -0.22) | <.001 | 884 | -0.21 (-0.30 to -0.12) | <.001 | 832 | -0.28 (-0.36 to -0.22) | < 0.001 |
| Grip strength | 919 | -0.05 (-0.09 to -0.01) | .017 | 913 | -0.10 (-0.15 to -0.04) | <.001 | 863 | -0.03 (-0.07 to 0.01) | 0.110 |
| Grooved pegboard | 901 | -0.27 (-0.33 to -0.20) | <.001 | 897 | -0.21 (-0.29 to -0.12) | <.001 | 845 | -0.27 (-0.34 to -0.21) | < 0.001 |
| Contrast sensitivity | 903 | -0.13 (-0.19 to -0.06) | <.001 | 899 | -0.10 (-0.19 to -0.01) | .036 | 847 | -0.11 (-0.18 to -0.04) | 0.002 |
| Audiometry: HF-PTA | 900 | 0.17 (0.10 to 0.23) | <.001 | 897 | 0.14 (0.05 to 0.23) | .003 | 845 | 0.19 (0.13 to 0.26) | < 0.001 |
| Audiometry: 4F-PTA | 901 | 0.20 (0.14 to 0.26) | <.001 | 897 | 0.14 (0.05 to 0.23) | .003 | 846 | 0.14 (0.07 to 0.21) | < 0.001 |
| LiSN-S Low cue | 901 | -0.17 (-0.23 to -0.10) | <.001 | 897 | -0.10 (-0.19 to -0.01) | .026 | 845 | -0.17 (-0.24 to -0.10) | < 0.001 |
| LiSN-S Spatial advantage | 901 | 0.22 (0.15 to 0.28) | <.001 | 897 | 0.18 (0.09 to 0.27) | <.001 | 845 | 0.21 (0.14 to 0.21) | < 0.001 |
| Physical limitations (SF-36) | 921 | 0.29 (0.23 to 0.35) | <.001 | 913 | 0.11 (0.02 to 0.19) | .012 | 864 | 0.29 (0.23 to 0.36) | < 0.001 |
| **Perceptions of aging** |  |  |  |  |  |  |  |  |  |
| Self-reported aging attitudes | 920 | -0.22 (-0.28 to -0.16) | <.001 | 911 | -0.25 (-0.34 to -0.16) | <.001 | 863 | -0.21 (-0.28 to -0.14) | < 0.001 |
| Self-reported health | 927 | -0.35 (-0.41 to -0.29) | <.001 | 916 | -0.27 (-0.36 to -0.19) | <.001 | 870 | -0.34 (-0.42 to -0.29) | < 0.001 |
| Self-reported perceived age | 892 | 0.09 (0.03 to 0.16) | .005 | 888 | 0.11 (0.01 to 0.20) | .024 | 838 | 0.08 (0.02 to 0.16) | 0.018 |
| Informant-reported health | 881 | -0.38 (-0.45 to -0.32) | <.001 | 875 | -0.30 (-0.38 to -0.21) | <.001 | 827 | -0.36 (-0.44 to -0.31) | < 0.001 |
| Researcher-reported health | 930 | -0.58 (-0.62 to -0.52) | <.001 | 916 | -0.45 (-0.51 to -0.37) | <.001 | 873 | -0.56 (-0.62 to -0.51) | < 0.001 |
| Informant-reported age appearance | 881 | 0.35 (0.29 to 0.41) | <.001 | 875 | 0.34 (0.25 to 0.43) | <.001 | 827 | 0.34 (0.29 to 0.42) | < 0.001 |
| Researcher-reported age appearance | 930 | 0.44 (0.38 to 0.49) | <.001 | 916 | 0.40 (0.31 to 0.47) | <.001 | 873 | 0.43 (0.37 to 0.50) | < 0.001 |
| Self-reported age appearance | 894 | 0.10 (0.03 to 0.16) | .003 | 891 | 0.11 (0.02 to 0.21) | .015 | 838 | 0.08 (0.01 to 0.15) | 0.029 |
| Facial age | 905 | 0.33 (0.26 to 0.39) | <.001 | 901 | 0.30 (0.22 to 0.39) | <.001 | 850 | 0.33 (0.28 to 0.41) | < 0.001 |
| Perceived Longevity | 908 | -0.27 (-0.33 to -0.20) | <.001 | 902 | -0.20 (-0.28 to 0.11) | <.001 | 852 | -0.26 (-0.32 to -0.19) | <0.001 |

Compared to peers with a slower Pace of Aging, those who were aging faster had slower gait speed (β = −0.33, p < 0.001; 95% CI: −0.39, −0.27), poorer balance (one-legged balance, β = 0.36, p < 0.001; 95% CI: −0.42, −0.30), were slower at rising repeatedly from a chair (chair stands, β = −0.30, p < 0.001; 95% CI: −0.37, −0.24) and stepping in place (two-minute step test, β = −0.28, p < 0.001; 95% CI: − 0.34, −0.22), were weaker (grip strength, β = −0.05, p = 0.017; 95% CI: −0.09, −0.01), and had more difficulties with fine motor control (grooved pegboard, β = −0.27, p < 0.001; 95% CI: −0.33, −0.20).

In addition, Study members who were aging faster had diminished sensory abilities. Visual contrast sensitivity and hearing ability are known to decline with advanced age.^34,38^ Study members with a faster Pace of Aging at age 45 had more difficulty visually distinguishing an object from its background on tests of contrast sensitivity (β = −0.13, p < 0.001; 95% CI: −0.19, −0.07). They also had more difficulties detecting high-pitch-tones (HF-PTA, β = 0.17, p < 0.001; 95% CI: 0.10, 0.23) and low-mid-pitch tones (4F-PTA, β = 0.20, p < 0.001; 95% CI: 0.14, 0.26) and were worse at hearing sentences in noisy environments when auditory distractors were nearby (LiSN-S Low cue, β = −0.17, p < 0.001; 95% CI: −0.23, −0.10) and when distractors were spatially distant (LiSN-S Spatial advantage, β = 0.22, p < 0.001; 95% CI: 0.15, 0.28). Finally, Study members with a faster Pace of Aging noticed sensory-motor difficulties in their everyday lives, self-reporting more physical limitations (SF-36 physical functioning scale, β = 0.29, p < 0.001; 95% CI: 0.23, 0.35). Sensitivity analyses revealed that associations between the Pace of Aging and sensory-motor difficulties were not attributable to being overweight, to smoking or to common age-related diseases (**Table 2**).

### Accelerated biological aging and negative perceptions of aging

Age-related morbidity and mortality are not only forecast by objective measures of physical and cognitive functioning. Older adults who self-report that they feel old are also more likely to subsequently be diagnosed with age-related disease and die at a younger age.^39,40^ We found that Study members with a faster Pace of Aging were more likely to hold unfavorable views of aging. **Table 2** reports effect sizes, significance tests and covariate-adjusted analyses for all perception measures.

Study members with a faster Pace of Aging had more negative attitudes towards aging (β = − 0.22, p < 0.001; 95% CI: −0.28, −0.16), endorsing sentiments such as “things keep getting worse as I get older” and “I am not as happy now as I was when I was younger.” They self-reported that they felt less healthy (β = −0.35, p < 0.001; 95% CI: −0.41, −0.29) and that they felt older than their chronological age (β = 0.09, p = 0.005; 95% CI: 0.03, 0.16). When asked similar questions about the Study members, informants (who knew them well) and research workers (who met the Study members during their one-day Unit visit) both reported that Study members with a faster Pace of Aging seemed in worse health (informant, β = −0.38, p < 0.001; 95% CI: −0.45, −0.32; research worker, β = −0.58, p < 0.001; 95% CI: −0.62, −0.52) and looked older than their age (informant, β = 0.35, p < 0.001; 95% CI: 0.29, 0.41; research worker, β = 0.44, p < 0.001; 95% CI: 0.38, 0.49; **Figure 4A**). In addition, Study members who were aging faster self-reported that they looked older than their age (β = 0.10, p = 0.003; 95% CI: 0.03, 0.16) and, when solely presented with facial images, independent raters scored Study members with a faster Pace of Aging as looking older than their peers (β = 0.33, p < 0.001; 95% CI: 0.26, 0.39) (**Figure 4B and 4C**). Finally, Study members with a faster Pace of Aging were less likely to think that they would live past the age of 75 (β = −0.27, p < 0.001; 95% CI: −0.33, −0.20). Sensitivity analyses revealed that associations between Pace of Aging and perceptions of aging were not attributable to being overweight, to smoking or to common age-related diseases (**Table 2**).

**Figure 4.**
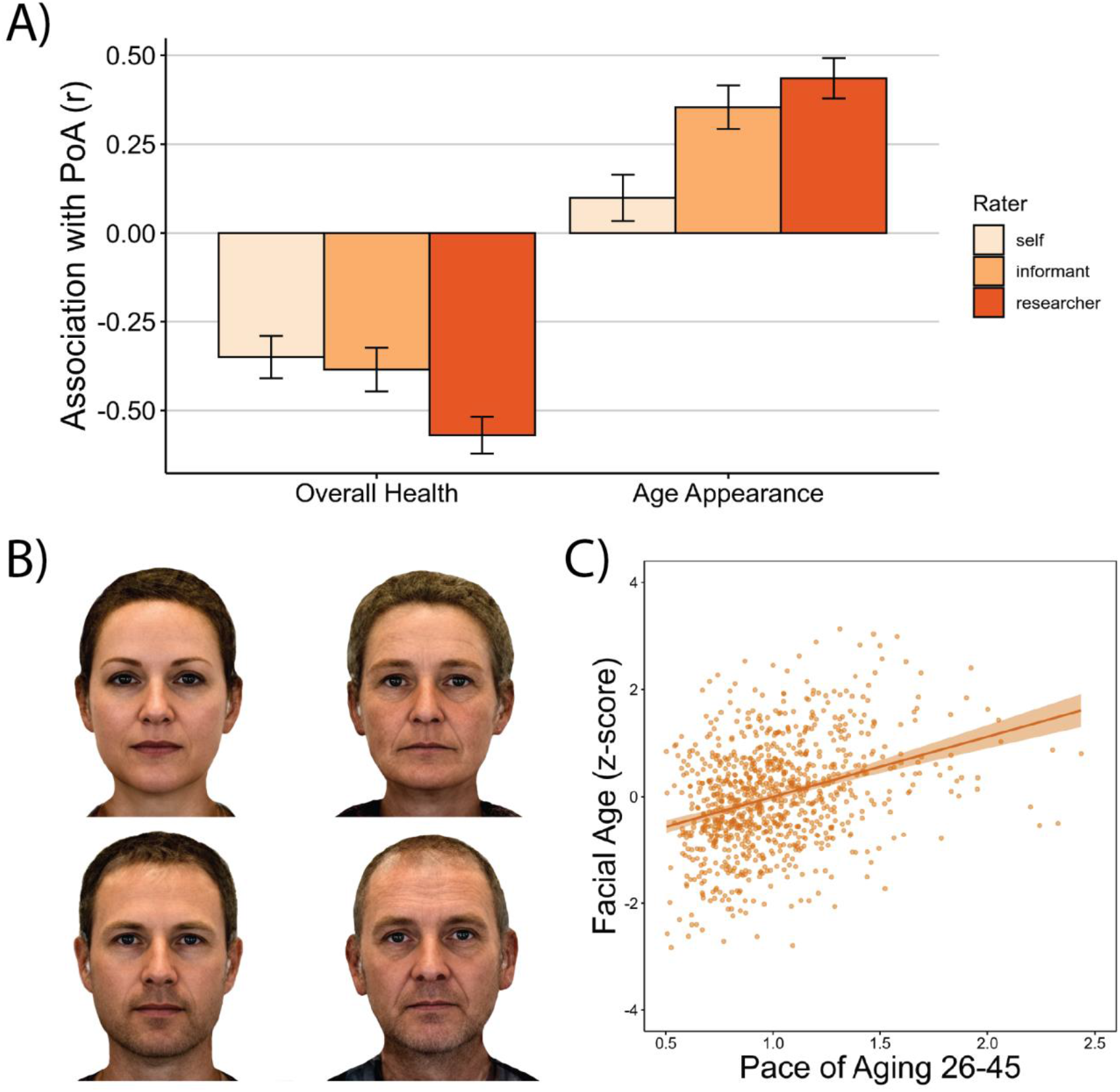
Study members who were aging faster were perceived as less healthy and looking older, when compared to slower-aging peers. (A) Effect sizes of associations between Pace of Aging and self-report, informant and research worker ratings of overall health and age appearance. (B) Digitally averaged composite faces made up of the ten male and female Study members with the youngest (left) and oldest (right) facial age ratings. (C) Scatterplot of the association between Pace of Aging and facial age ratings by independent raters. Scatterplot includes a regression line with 95% confidence interval.

## DISCUSSION

Chronological age is a poor proxy for biological age, even in midlife. Here, in a population-representative birth cohort without variation in chronological age, we found that Study members had large variation in their Pace of Aging. Furthermore, Study members who had a faster Pace of Aging in midlife exhibited signs of advanced brain aging, experienced more cognitive difficulties, had diminished sensory-motor functional capacity and had more negative perceptions of aging. Sensitivity analyses revealed that these associations were not explained by obesity, smoking or chronic diseases, supporting our hypothesis that the Pace of Aging is a robust indicator of the cumulative, progressive, and gradual deterioration across organ systems that underlies biological aging. Together, these findings support at least two conclusions: 1) meaningful variation in biological aging can be measured in midlife; and 2) people with a faster rate of biological aging across the first half of the lifespan are more likely to experience age-related functional impairment by midlife. These findings raise the question of whether midlife is a window of opportunity for the mitigation of age-related disease. We have shown that biological aging in midlife is meaningful, yet further research is needed to determine whether biological aging in midlife is still malleable. Randomized trials are beginning to test this possibility.^41,42^

Four design features of the Dunedin Study support these conclusions. First, all Study members were born in 1972-1973, which allows the direct measurement of individual differences in biological aging uncoupled from age and cohort effects.^43,44^ Second, the Dunedin Study has very low attrition rates; unlike many longitudinal studies of older adults that have selective attrition and mortality, the full range of health is represented.^45,46^ Third, the Dunedin Study has collected four waves of biological measurements from age 26 to age 45, a unique dataset allowing for more accurate estimates of biological aging. Fourth, even though age-related diseases are uncommon in midlife, Study members were assessed at age 45 with a battery of established measures that are commonly used in geriatric settings to predict frailty, morbidity and mortality.

This study was not without limitations. First, these findings are based on a single birth cohort from New Zealand. Second, our study currently lacks follow-up past the age of 45. Further investigation of the Pace of Aging in diverse cohorts and in older adults is needed. Third, the Pace of Aging was derived from 19 biomarkers repeatedly assessed across 20 years, which will be infeasible for most studies of biological aging. However, we recently reported that a proxy for the Pace of Aging can now be quantified from genome-wide DNA methylation data extracted from a single cross-sectional blood draw.^47^ This measure makes it possible for studies lacking 4 waves of biomarkers to extend this work; e.g., it predicts disease and mortality in U.S. and U.K. samples.^47^ Fourth, while associations were consistent across domains and measures, effect-sizes were generally moderate. However, these moderate associations between the Pace of Aging and midlife function likely reflect the cumulative effects of the aging process. Therefore, if the Pace of Aging truly measures the underlying aging process, the associations reported here should grow larger over time, as fast and slow agers continue to diverge.^48^

Within the bounds of these limitations, our findings have implications for geroscience theory, research, and policy. With regard to theory, we find that variation in the pace of biological aging can be quantified in midlife and, among 45-year-olds, is already associated with indicators of risk for frailty, morbidity and mortality across 4 domains. The breadth of these associations is consistent with the geroscience hypothesis depicting accelerated aging as a common cause of age-related chronic disease. While previous research suggested that biological aging could be measured in younger adults,^22^ it was unclear whether these measures indexed biological aging with functional implications for later-life health. Here we show that faster aging in midlife is associated with several measures of functional impairment and frailty that in older adults have established links to morbidity and mortality. For example, Study members who were in the fastest quintile of the Pace of Aging had brainAGEs that were an average of 3.79 years older and were rated as looking 4.32 years older than those in the slowest quintile. Further research is needed to test whether interventions in humans can slow biological aging in midlife and reduce long-term risk for age-related chronic disease. Interventions that can achieve even mild slowing of biological aging promise to improve quality of life in older adults while yielding significant healthcare savings.^17,49^

With regard to research and policy, current efforts aimed at improving biological aging measurement are primarily driven by the need to test emerging anti-aging biotechnology. While our findings support these efforts, they also suggest that biological aging may have broader implications for society. Many social programs, including state pensions and Medicare in the U.S., are designed to offset the economic and health burdens that accrue as individuals age. Eligibility for these benefits has historically been determined on the basis of chronological age. For example, the age for U.S. Social Security eligibility was set to 65 in 1939 when the average life expectancy was 63.7.^50^ However, with lengthening lifespans, it is important to also consider biological age. Our findings suggest that already by midlife, chronological age is a crude, poorly calibrated measure of the functional consequences of aging. We provide evidence that disparities in biological aging independent of chronological age are already linked to functional difficulties in midlife. Some of those difficulties may reduce individuals’ ability to work. Widespread application of biological aging measures could represent an alternative to using birthdates when determining the allocation of healthcare and financial support for those suffering from the sequelae of aging. For example, in the U.S., there are ongoing debates about lowering the Medicare age to expand access to preventative healthcare.^51,52^ Perhaps someday we will be able to use biological aging measures to guide treatment access. With further development, geroscience could provide the conceptual tools, measurement technology, and interventions required to mitigate disparities in the pace of biological aging through more tailored and just access to independence-sustaining resources.

## METHODS

### Study Design and Population

Participants are members of the Dunedin Study, a longitudinal investigation of health and behavior in a representative birth cohort. The 1037 participants (91% of eligible births) were all individuals born between April 1972 and March 1973 in Dunedin, New Zealand, who were eligible on the basis of residence in the province and who participated in the first assessment at age 3 years^21^. The cohort represents the full range of socioeconomic status (SES) in the general population of New Zealand’s South Island and, as adults, matches the New Zealand National Health and Nutrition Survey on key adult health indicators (e.g., body mass index, smoking, and general practitioner visits) and the New Zealand Census of citizens of the same age on educational attainment^21,53^. The cohort is primarily white (93%, self-identified), matching South Island demographic characteristics. Assessments were performed at birth; at ages 3, 5, 7, 9, 11, 13, 15, 18, 21, 26, 32, and 38 years; and, most recently (completed April 2019), at age 45 years, when 938 of the 997 participants (94.1%) still alive participated. Study members with data available at age 45 years did not differ significantly from other living participants in terms of childhood SES or childhood neurocognitive functioning (see attrition analysis in **Supplemental Figure S1 and S2**). At each assessment, each participant was brought to the research unit for interviews and examinations. Research staff make standardized ratings, informant questionnaires are collected, and administrative records are searched. Written informed consent was obtained from cohort participants. Ethical approval for the study protocols was given by the campus Institutional Review Board of Duke University and the Health and Disability Ethics Committees of the New Zealand government. This study follows the Strengthening the Reporting of Observational Studies in Epidemiology (STROBE) reporting guideline. The premise and analysis plan for this project were pre-registered at https://bit.ly/2ZVtnsq.

### Statistical Analysis

Unless otherwise specified, all statistical analyses were completed using linear regression models in R (version 3.4.0). All models were adjusted for sex. In addition, two types of sensitivity analyses were also performed for all associations. 1) In addition to sex, body mass index and smoking status at age 45 were simultaneously added as covariates to rule out the possibility that associations were limited to overweight individuals and to smokers. 2) Sex-adjusted models were run in which all Study members were excluded who had diagnosed, common age-related diseases (cancer, diabetes, heart attack). Correction for multiple comparisons was performed using the false discovery rate correction across all 38 sex-adjusted models presented in Table 1 and Table 2. Analyses reported here were checked for reproducibility by an independent data-analyst who recreated the code by working from the manuscript and applied it to an independently generated copy of the dataset.

### Measuring the Pace of Aging

#### Pace of Aging

We measured Pace of Aging from repeated assessments of a panel of 19 biomarkers: Body mass index (BMI), Waist-hip ratio, Glycated hemoglobin, Leptin, Blood pressure (mean arterial pressure), Cardiorespiratory fitness (VO_2_Max), Forced vital capacity ratio (FEV_1_/FVC), Forced expiratory volume in one second (FEV_1_), Total cholesterol, Triglycerides, High density lipoprotein (HDL), Lipoprotein(a), Apolipoprotein B100/A1 ratio, estimated Glomerular Filtration Rate (eGFR), Blood Urea Nitrogen (BUN), High Sensitivity C-reactive Protein (hs-CRP), White blood cell count, mean periodontal attachment loss (AL), and the number of dental-caries-affected tooth surfaces (tooth decay). Biomarkers were assayed at the age-26, 32, 38 and 45 assessments. The Pace of Aging reported here represents an extension of a previously reported measure that used 18 biomarkers assayed at ages 26, 32 and 38.^22^ Here we add a recently completed 4^th^ measurement wave of data, at age 45, totaling 19 biomarkers. We added measures of leptin and carries, both of which have now been assessed at multiple waves allowing growth curve modeling. Telomere length was dropped because of an emerging and yet-unresolved field-wide debate about its measurement^54^. Details on biomarker measurements are provided in **Supplemental Table S1**.

We calculated each Study member’s Pace of Aging in three steps. In the first step, we transformed the biomarker values to a standardized scale. For each biomarker at each wave, we standardized values according to the age-26 distribution (i.e. set to mean of 0 and a standard deviation of 1). Standardization was conducted separately for men and women. Standardized biomarker values greater than zero indicated levels that were “older” and values less than zero indicated levels “younger” than the average 26-year-old. To match, scores were reversed for VO_2_Max, FEV_1_/FVC, FEV_1_, eGFR, and HDL cholesterol, which are known to decline with age. Over the 2 decades of follow-up, the biomarkers in the panel indicated a progressive deterioration of physiological integrity with advancing chronological age; i.e. their cohort mean values tended to increase (i.e., worsen) from the age-26 assessment to the age-45 assessment (**Figure 2**).

In the second step, we calculated each Study member’s personal slope for each of the 19 biomarkers—the average year-on-year change observed over the 2-decade period. Slopes were estimated using a mixed effects growth model that regressed the biomarker’s level on age. A complete list of means of biomarker slopes and pairwise correlations among biomarker slopes is presented in **Supplemental Table S2**. For only four of the 19 biomarkers we examined, cohort mean levels did not worsen over time as expected based on published associations with age-related chronic disease: white blood cell count and CRP levels remained stable with age; HDL cholesterol and apolipoprotein B100/A1 ratio improved with age. However, individual-difference slopes for these biomarkers did show the expected pattern of correlation with other biomarkers’ slopes. For example, Study members whose apolipoprotein B100/A1 ratio increased during the follow-up period also showed increasing adiposity, declining lung function, and increasing systemic inflammation. We retained all pre-registered biomarkers in the Pace of Aging model.

In the third step, we combined information from the 19 slopes of the biomarkers to calculate each Study member’s personal “Pace of Aging.” Because we did not have any *a priori* basis for weighting differential contributions of the biomarkers to an overall Pace of Aging measure, we combined information using a unit-weighting scheme (all biomarkers were standardized to have mean=0, SD=1 based on their age-26 distributions, so slopes were denominated in comparable units). We calculated each Study member’s Pace of Aging as the sum of age-dependent annual changes in biomarker Z-scores. Because the Dunedin birth cohort represents its population, its mean and distribution represent population norms. We used these norms to scale the Pace of Aging to reflect physiological change relative to the passage of time. We set the cohort mean Pace of Aging as a reference value equivalent to the physiological change expected during a single chronological year. Using this reference value, we rescaled Pace of Aging in terms of years of physiological change per chronological year (M = 1, SD = 0.29). On this scale, cohort members ranged in their Pace of Aging from 0.4 years of physiological change per chronological year (slow) to 2.4 years of physiological change per chronological year (fast) (**Figure 2**).

As a sensitivity check to ensure that the geroscience definition of aging as unidirectional decline fits the data, we examined biomarker patterns of change for potential non-linearity. Three biomarkers – leptin, hs-CRP and eGFR – were measured at only three time-points and could only be fit with a linear model. For all other biomarkers, we fit an additional model that included fixed effects for the intercept, linear change and quadratic change, as well as random effects for the intercept and linear terms. For nine biomarkers, fit statistics (residual LL, AIC, BIC) indicated that the linear model provided a better fit than the quadratic model. For seven biomarkers, fit statistics indicated that the quadratic model provided a better fit than the linear model. However, for these seven biomarkers, the linear slope estimates extracted from the two models were highly correlated in sex-adjusted models (Waist-hip ratio: 0.99, VO_2_Max: 1.00, FEV_1_/FVC: 0.99, FEV_1_: 0.99, Apolipoprotein B100/A1 ratio: 0.99; BUN: 0.99; Gum health: 0.99), leading us to conclude that we could reasonably use the linear slope estimates from the models including linear fixed effects only. This is graphically apparent in **Supplemental Figure S3**, which compares the linear-only and linear + quadratic growth curves.

### Structural MRI

#### Image Acquisition

Each participant was scanned using a Siemens MAGNETOM Skyra (Siemens Healthcare GmbH) 3T scanner equipped with a 64-channel head/neck coil at the Pacific Radiology Group imaging center in Dunedin, New Zealand. High resolution T1-weighted images were obtained using an MP-RAGE sequence with the following parameters: TR = 2400 ms; TE = 1.98 ms; 208 sagittal slices; flip angle, 9°; FOV, 224 mm; matrix =256×256; slice thickness = 0.9 mm with no gap (voxel size 0.9×0.875×0.875 mm); and total scan time = 6 min and 52 s. 3D fluid-attenuated inversion recovery (FLAIR) images were obtained with the following parameters: TR = 8000 ms; TE = 399 ms; 160 sagittal slices; FOV = 240 mm; matrix = 232×256; slice thickness = 1.2 mm (voxel size 0.9×0.9×1.2 mm); and total scan time = 5 min and 38 s. Additionally, a gradient echo field map was acquired with the following parameters: TR = 712 ms; TE = 4.92 and 7.38 ms; 72 axial slices; FOV = 200 mm; matrix = 100×100; slice thickness = 2.0 mm (voxel size 2 mm isotropic); and total scan time = 2 min and 25 s. Diffusion-weighted images providing full brain coverage were acquired with 2.5 mm isotropic resolution and 64 diffusion weighted directions (4700 ms repetition time, 110.0 ms echo time, b value 3,000 s/mm2, 240 mm field of view, 96×96 acquisition matrix, slice thickness=2.5 mm). Non-weighted (b=0) images were acquired in both the encoding (AP) and reverse encoding (PA) directions to allow for EPI distortion correction. 875 Study members completed the MRI scanning protocol (see **Supplemental Figures S1 and S2** for attrition analyses).

#### Image Processing

Structural MRI data were analyzed using the Human Connectome Project (HCP) minimal preprocessing pipeline as detailed elsewhere.^55^ Briefly, T1-weighted and FLAIR images were processed through the PreFreeSurfer, FreeSurfer, and PostFreeSurfer pipelines. T1-weighted and FLAIR images were corrected for readout distortion using the gradient echo field map, coregistered, brain-extracted, and aligned together in the native T1 space using boundary-based registration^56^. Images were then processed with a custom FreeSurfer recon-all pipeline that is optimized for structural MRI with higher resolution than 1 mm isotropic. Finally, recon-all output were converted into CIFTI format and registered to common 32k_FS_LR mesh using MSM-sulc.^57^ Outputs of the minimal preprocessing pipeline were visually checked for accurate surface generation by examining each participant’s myelin map, pial surface, and white matter boundaries.

#### Cortical thickness, surface area and hippocampal volume

For each participant the mean cortical thickness and surface area were extracted from each of the 360 cortical areas in the HCP-MPP1.0 parcellation.^58^ Regional cortical thickness and surface area measures have each been found to have excellent test-retest reliability in this sample (mean ICCs = 0.85 and 0.99 respectively).^59^ Bilateral hippocampal volume was extracted from the FreeSurfer “aseg” parcellation. Of the 875 Study members for whom data were available, 4 were excluded due to major incidental findings or previous injuries (e.g., large tumors or extensive damage to the brain/skull), 9 due to missing FLAIR or field map scans, 1 due to poor surface mapping yielding 861 participants’ datasets for cortical thickness surface area and hippocampal volume analyses.

#### White matter hyperintensities

To identify and extract the total volume of white matter hyperintensities (WMH), T1-weighted and FLAIR images for each participant were processed with the UBO Detector, a cluster-based, fully-automated pipeline with established out-of-sample performance, and high reliability in our data (test-retest ICC = 0.87).^60,61^ The resulting WMH probability maps were thresholded at 0.7, which is the suggested standard. WMH volume is measured in Montreal Neurological Institute (MNI) space, thus removing the influence of differences in brain volume on WMH volume. Because of the potential for bias and false positives due to the thresholds and masks applied in UBO, the resulting WMH maps for each participant were manually checked by two independent raters to ensure that false detections did not substantially contribute to estimates of WMH volume. Visual inspections were done blind to the participants’ cognitive status. Due to the tendency of automated algorithms to mislabel regions surrounding the septum as WMH, these regions were manually masked out to further ensure the most accurate grading possible. For WMH data, participants were excluded if they had missing FLAIR scans, Multiple Sclerosis, or inaccurate white matter labelling or low-quality MRI data, yielding 852 participants’ datasets for analyses. In all analyses, WMH volume was log-transformed.

#### Diffusion Weighted Imaging

Diffusion weighted images were processed in FSL (http://fsl.fmrib.ox.ac.uk/fsl). Raw diffusion weighted images were corrected for susceptibility artifacts, subject movement, and eddy currents using topup and eddy. Images were then skull-stripped and fitted with diffusion tensor models at each voxel using FMRIB’s Diffusion Toolbox (FDT; http://fsl.fmrib.ox.ac.uk/fsl/fslwiki/FDT). The resulting fractional anisotropy (FA) images from all Study members were non-linearly registered to the FA template developed by the Enhancing Neuro Imaging Genetics Through Meta-Analysis consortium (ENIGMA), a minimal deformation target calculated across a large number of individuals.^62^ The images were then processed using the tract-based spatial statistics (TBSS) analytic method^63^ modified to project individual FA values onto the ENIGMA-DTI skeleton. Following the extraction of the skeletonized white matter and projection of individual FA values, ENIGMA-tract-wise regions of interest, derived from the Johns Hopkins University (JHU) white matter parcellation atlas,^64^ were transferred to extract the mean FA across the full skeleton and average FA values for a total of 25 (partially overlapping) regions. After visual inspection of all diffusion images, 7 Study members were removed because data were collected with 20-channel head coils, leading to poor diffusion image quality; 3 were removed due to major incidental findings; 5 were removed due to excessive (>3mm) motion detected with eddy tool; and 7 were removed due to missing diffusion scans. This resulted in 854 Study members with high-quality diffusion images that were included in the analysis.

#### Brain age

We estimated brain age with a publicly available algorithm, developed by a different research team, which uses information about cortical anatomy to estimate the age of a person’s brain.^31^ They trained this algorithm on chronological age in samples ranging from 19-82 years old. The algorithm has been shown to predict chronological age in multiple independent samples, and to have high test-retest reliability in the Dunedin Study (ICC = 0.81),^65^ although it has a documented tendency to underestimate chronological age by approximately 3 years among adults between chronological ages 44 and 46 years. For this reason, we standardized the scores to the mean chronological age of the Dunedin Study members at the time of their scanning in the Phase-45 assessment.^66^ In all analyses we used the brain Age Gap Estimate or brainAGE, which is the difference between each Study member’s estimated brain age and their chronological age. An older brainAGE results when the predicted brain age is older than the study member’s chronological age and is presumed to reflect accelerated brain aging. Data from six Study members were excluded due to major incidental findings or previous head injuries (e.g., large tumors or extensive damage to the brain). This resulted in brainAGE data from 869 Study members.

### Cognitive functioning

#### Neurocognitive functioning

The Wechsler Adult Intelligence Scale-IV (WAIS-IV)^67^ was administered to each participant at age 45 years, yielding the IQ. In addition to full scale IQ, the WAIS-IV yields indexes of four specific cognitive function domains: Processing Speed, Working Memory, Perceptual Reasoning, and Verbal Comprehension.

#### Child-to-adult neurocognitive decline

The Wechsler Intelligence Scale for Children–Revised (WISC–R)^68^ was administered to each participants at ages 7, 9, and 11 years, yielding the IQ. To increase baseline reliability, we averaged each participant’s three scores. We measured cognitive decline by studying IQ scores at midlife after controlling for IQ scores in childhood (as a sensitivity analysis, in addition to analyzing residualized change, we also analyzed “change scores” assessed as the difference between adult IQ and childhood IQ, and obtained the same substantive and statistically-significant results). We focus on change in the overall IQ given evidence that age-related slopes are correlated across all cognitive functions, indicating that research on cognitive decline may be best focused on a highly reliable summary index, rather than focused on individual functions^69^.

#### Rey Auditory Verbal Learning Test

This is a test of verbal learning and memory administered at 45 years.^70^ The test involves repeated presentation of a 15-word list and a one-time presentation of an interference list. Total Recall is the total number of words (0-60) recalled over four trials (the sum of words recalled across trials 1-4). Delayed Recall is the total number of words (0-15) recalled after a 30-minute delay.

#### Informant memory and attention

Subjective everyday cognitive function was reported by individuals nominated by each participant as knowing him/her well. These informants were mailed questionnaires and asked to complete a checklist indicating whether the Study member had problems with memory or attention over the past year. 94% of Study members had at least one informant return the questionnaire, 88% had two, and 68% had three. A memory-problems scale consisted of three items: “has problems with memory,” “misplaces wallet, keys, eyeglasses, paperwork,” and “forgets to do errands, return calls, pay bills” (internal consistency reliability = 0.63). An attention-problems scale consisted of four items: “is easily distracted, gets sidetracked easily,” “can’t concentrate, mind wanders,” “tunes out instead of focusing,” and “has difficulty organizing tasks that have many steps” (internal consistency reliability = 0.79).

#### Sensory-motor Functioning

We assessed sensory-motor functional capacity at age 45 with objective tests of physical and sensory functioning and self-reports of physical limitations.

##### Gait speed

Gait speed (meters per second) was assessed with the 6-m-long GAITRite Electronic Walkway (CIR Systems, Inc) with 2-m acceleration and 2-m deceleration before and after the walkway, respectively. Gait speed was assessed under 3 walking conditions: usual gait speed (walk at normal pace from a standing start, measured as a mean of 2 walks) and 2 challenge paradigms, dual-task gait speed (walk at normal pace while reciting alternate letters of the alphabet out loud, starting with the letter “A,” measured as a mean of 2 walks) and maximum gait speed (walk as fast as safely possible, measured as a mean of 3 walks). Gait speed was correlated across the 3 walk conditions.^71^ To increase reliability and take advantage of the variation in all 3 walk conditions (usual gait and the 2 challenge paradigms), we calculated the mean of the 3 highly correlated individual walk conditions to generate our primary measure of composite gait speed.

#### One-legged balance

Balance was measured using the Unipedal Stance Test as the maximum time achieved across three trials of the test with eyes closed.^72–74^

#### Chair-stand test

Chair rises were measured as the number of stands with no hands a participant completed in 30 seconds from a seated position.^75,76^

#### 2 min step test

The 2-min step test was measured as the number of times a participant lifted their right knee to mid-thigh height (measured as the height half-way between the knee cap and the iliac crest) in 2 minutes at a self-directed pace.^76,77^

#### Grip strength

Handgrip strength was measured for each hand (elbow held at 90°, upper arm held tight against the trunk) as the maximum value achieved across three trials using a Jamar digital dynamometer.^36,78^

#### Visual-motor coordination

Visual-motor coordination was measured as the time to completion of the Grooved Pegboard Test.^70^ Scores for the Grooved Pegboard test were reversed so that higher values corresponded to better performance.

#### Contrast Sensitivity

Study members wore their glasses or contact lenses (if these were normally worn). Study members were seated one meter from the Thomson Test Chart and the Samsung 23” LCD Thin Client screen. Room lighting was set at 520 lux. Contrast sensitivity was tested with both eyes open. The Pelli-Robson chart presents three letters per line and the black letters gradually fade from black to grey to white on the white background to determine the lowest level of “contrast” that the eye can detect. If only one letter on a line was correctly determined by the study member, the number of letters was recorded to determine the CSF score. However, if two letters on a line were correctly determined, the technician proceeded to the next line to determine if the study member could correctly determine any of these letters.

#### Audiometry

Hearing acuity was assessed in a sound-attenuating booth (350 Series MaxiAudiology Booth by IAC Acoustics) which met the standard for maximum permissible ambient sound pressure levels. Pure tone audiometry was administered via the Interacoustics Callisto Suite configured to the Interacoustics OtoAccess database, operated from an HP Envy laptop with sound delivered by Sennheiser HDA 300 headphones. The program was set to deliver pure-tone stimuli in the following order: 1000 Hz, 2000 Hz, 4000 Hz, 8000 Hz, 12500 Hz, and 500 Hz. Presentation intensity levels began at 40 decibels at hearing level (dB HL) for normal hearing Study members, and 60 dB HL for hearing aid users. Audiometry used the Hughson-Westlake procedure (ISO8253-1:2010; Acoustics-Audiometric test methods-Part1: Pure-tone air and bone-conduction audiometry) in which participants respond when they hear a pure tone. Auditory thresholds, defined as the lowest intensity level that the individual responded to, for 2 out of 3 presentations, were determined using a standard down-10-up-5 technique for each frequency. A four-frequency pure-tone average was calculated by averaging 500 Hz, 1000 Hz, 2000 Hz, and 4000 Hz; and a high pure-tone average was calculated by averaging 8000 Hz and 12500 Hz. The results for the “best ear” are reported.

#### Spatial listening

Study members completed the Listening in Spatialised Noise–Sentences Test (LiSN-S) (Phonak, Switzerland) in a sound-attenuating booth (350 Series MaxiAudiology Booth by IAC Acoustics). Auditory stimuli were delivered through a pair of Sennheiser 215 headphones attached to a Mini PCM2704 external sound card. The LiSN-S produces a three-dimensional auditory environment through the headphones via four different task conditions.^79^ Target sentences are superimposed with distractor stories (maskers). Across the four conditions, these maskers differ with respect to perceived spatial location (0° or ±90 ° azimuth), and speaker identity (same or different to the target speaker). The following order of conditions was identically presented to all participants: 1) different speaker at ±90 ° azimuth, 2) same speaker at ±90 ° azimuth, 3) different speaker at ±0 ° azimuth, and 4) same speaker at ±0 ° azimuth.

The masking stories were consistently presented at an intensity of 55 decibels sound pressure level (dB SPL). Participants repeated the target sentences and were scored in the software on their accuracy (words correct in each sentence). The program was adaptive, with target sentences delivered at 62 dB SPL to start, and intensity levels continuously adjusted up (if <50% of the words in the sentence correct), and down (if >50% of the words in the sentence correct), based on accuracy. The first few sentences (a minimum of 5) are considered practice sentences. This practice testing continues where levels were lowered in 4 dB increments, until one upward reversal in performance was recorded (i.e. the sentence score drops <50% of words correct), after which the increments decreased to ±2 dB steps. Practice sentence scores did not form part of the final score. The test condition continued until the average of the levels from positive-and negative-going reversals amounted to ≥3 (independent midpoint target level), and the standard error of these midpoints was less than 1 dB. Alternatively, the test condition continued until it reached the maximum number of 30 sentence presentations. Speech-reception thresholds were calculated as the lowest intensity at which the individual could repeat 50% of the words correctly. Two outcome scores were used: 1) Speech reception threshold from a low-cue condition represented performance in the most difficult auditory environment (masker speaker same as the target speaker, and masker was presented at 0 ° azimuth, in the same location as the target speaker). 2) “Spatial advantage” score measured the benefit gained when the masker is presented from a different direction to the target.

#### Physical limitations

Physical limitations were measured with the RAND 36-Item Health Survey 1.0 physical functioning scale.^80^ Participant responses (“limited a lot”, “limited a little”, “not limited at all”) assessed their difficulty with completing various activities, e.g., climbing several flights of stairs, walking more than 1 km, participating in strenuous sports, etc. Scores were reversed to reflect physical limitations so that a high score indicates more limitations.

### Perceptions of aging

#### Attitudes towards Aging

Age beliefs were assessed with the five-item Attitude Toward Aging scale.^40^ Sample items: “Things keep getting worse as I get older (R)”, “As you get older, you are less useful.”

#### Perceived Health

We obtained three reports about Study members’ health from three sources: self-reports, informant impressions, and staff impressions (see next paragraph for a description of these data sources). All reporters rated the study member’s general health using the following response options: excellent, very good, good, fair or poor. Correlations between self-, informant-, and staff-ratings ranged from 0.48-0.55.

#### Age appearance

We obtained reports about Study members’ age appearance from three sources: self-reports, informant impressions, and staff impressions. *Self-reports* – We asked the Study members about their own impressions of how old they looked, “Do you think you LOOK older, younger or about your actual age?” Response options were younger than their age, about their actual age, or older than their age. We also asked Study members to rate their age perceptions in years, “How old do you feel?”. *Informant impressions* - Informants who knew a Study member well (94% response rate) were asked: “Compared to others their age, do you think he/she (the study member) looks younger or older than others their age? Response options were: much younger, a bit younger, about the same, a bit older, or much older. *Staff impressions* - Four members of Dunedin Study Unit staff competed a brief questionnaire describing each study member. To assess age appearance, staff used a 7-item scale to assign a “relative age” to each study member (1=young looking”, 7=“old looking). Correlations between self-, informant-, and staff-ratings ranged from 0.34-0.52.

#### Facial Age

Facial Age was based on ratings by an independent panel of eight raters of each participant’s digital facial photograph. Facial Age was based on two measurements of perceived age. First, Age Range was assessed by an independent panel of four raters, who were presented with standardized (non-smiling) facial photographs of participants and were kept blind to their actual age. Raters used a Likert scale to categorize each participant into a 5-year age range (i.e., from 20-24 years old up to 70+ years old) (interrater reliability = 0.77). Scores for each participant were averaged across all raters. Second, Relative Age was assessed by a different panel of four raters, who were told that all photos were of people aged 45 years old. Raters then used a 7-item Likert scale to assign a “relative age” to each participant (1=“young looking”, 7=“old looking”) (interrater reliability = .79). The measure of perceived age at 45 years, Facial Age, was derived by standardizing and averaging Age Range and Relative Age scores.

#### Perceived Longevity

At age 45, study members were asked, “How likely is it that you will live to be 75 or more?” (0=not likely, 1=somewhat likely, 2=very likely).

## Data Availability

The Dunedin Study datasets reported in the current article are not publicly available due to lack of informed consent and ethical approval, but are available on request by qualified scientists. Requests require a concept paper describing the purpose of data access, ethical approval at the applicant's university and provision for secure data access (https://moffittcaspi.trinity.duke.edu/research-topics/dunedin). We offer secure access on the Duke, Otago and King's College campuses.

## Data and Code Availability

Dunedin study data are available via managed data access (https://moffittcaspi.trinity.duke.edu/research). All code used this analysis are available upon request.

## Acknowledgements

This research was supported by National Institute on Aging (NIA) grants R01AG032282 and R01AG049789, and UK Medical Research Council grant MR/P005918/1. Additional support was provided by the Jacobs Foundation, NIA P30 AG028716 and NIA P30 AG034424. The Dunedin Multidisciplinary Health and Development Research Unit was supported by the New Zealand Health Research Council (Project Grants 15-265 and 16-604) and New Zealand Ministry of Business, Innovation, and Employment (MBIE). MLE is supported by the National Science Foundation Graduate Research Fellowship (NSF DGE-1644868). We thank members of the Advisory Board for the Dunedin Neuroimaging Study, Dunedin Study members, Unit research staff, Pacific Radiology Group staff, and Study founder Phil Silva, PhD.

