## Supplemental for "Disparities in the pace of biological aging among midlife adults of the same chronological age have implications for future frailty risk and policy"

**Supplemental Figure S1.** Childhood IQ attrition analysis.
**Supplemental Figure S2.** Childhood SES attrition analysis.
**Supplemental Figure S3.** Testing biomarker change for nonlinearity.
**Supplemental Table S1.** Measurement of biomarkers used to calculate Pace of Aging measures.
**Supplemental Table S2.** Pairwise correlations among Study-member-specific slopes for 19 biomarkers.
**Supplemental Table S3.** Parcel-wise cortical thickness results.
**Supplemental Table S4.** Parcel-wise surface area results.

**Supplemental Figure S1**. **Childhood IQ attrition analysis.** No significant differences in childhood IQ were found between the full cohort, those still alive, those seen at Phase 45 or those MRI-scanned at Phase 45. Those who were deceased by the Phase 45 data collection had significantly lower childhood IQ's than those who were still alive (t = 2.09, p = 0.04).

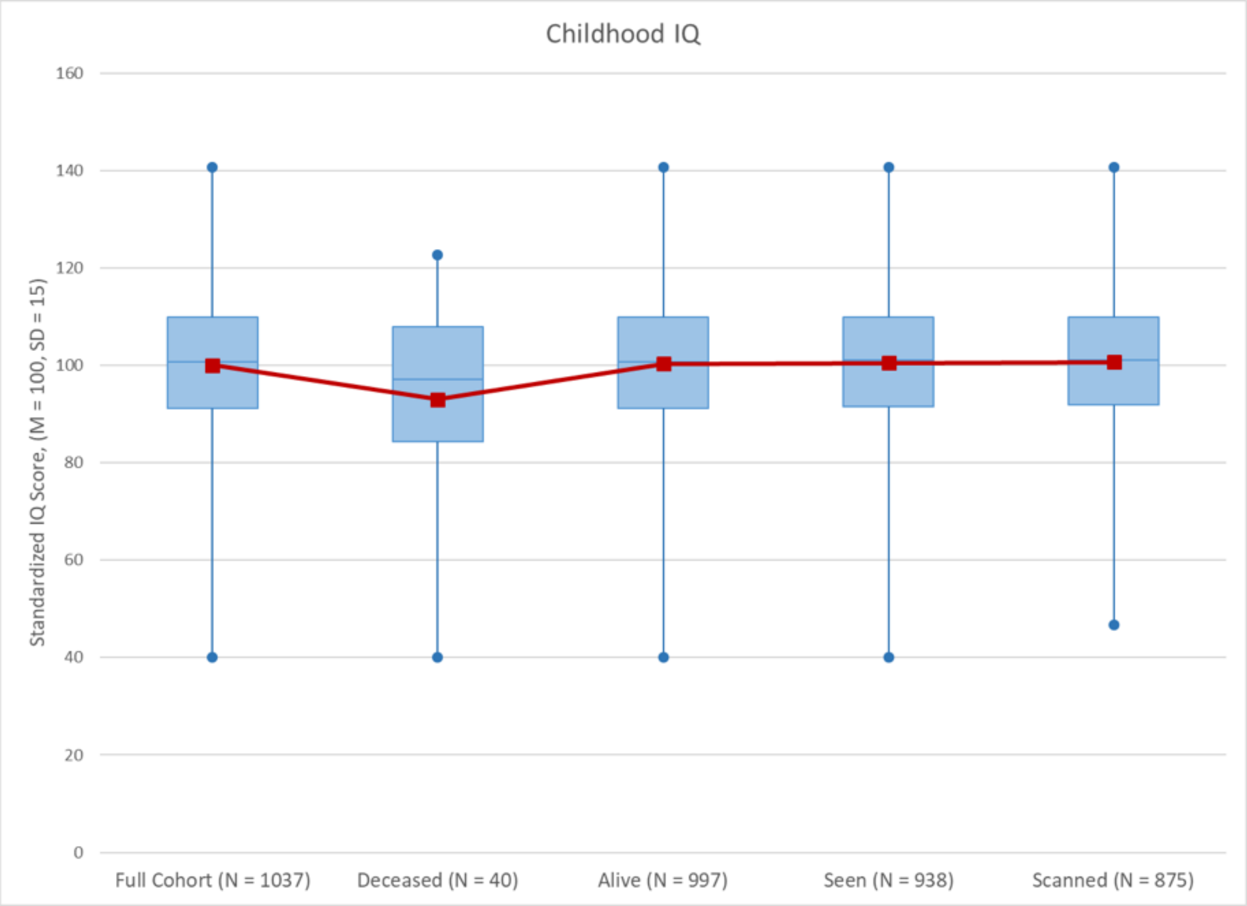

**Supplemental Figure S2. Childhood SES attrition analysis.** No significant differences were found between the full cohort, those deceased, those alive, those seen at Phase 45 or those MRI-scanned at Phase 45 on childhood SES.

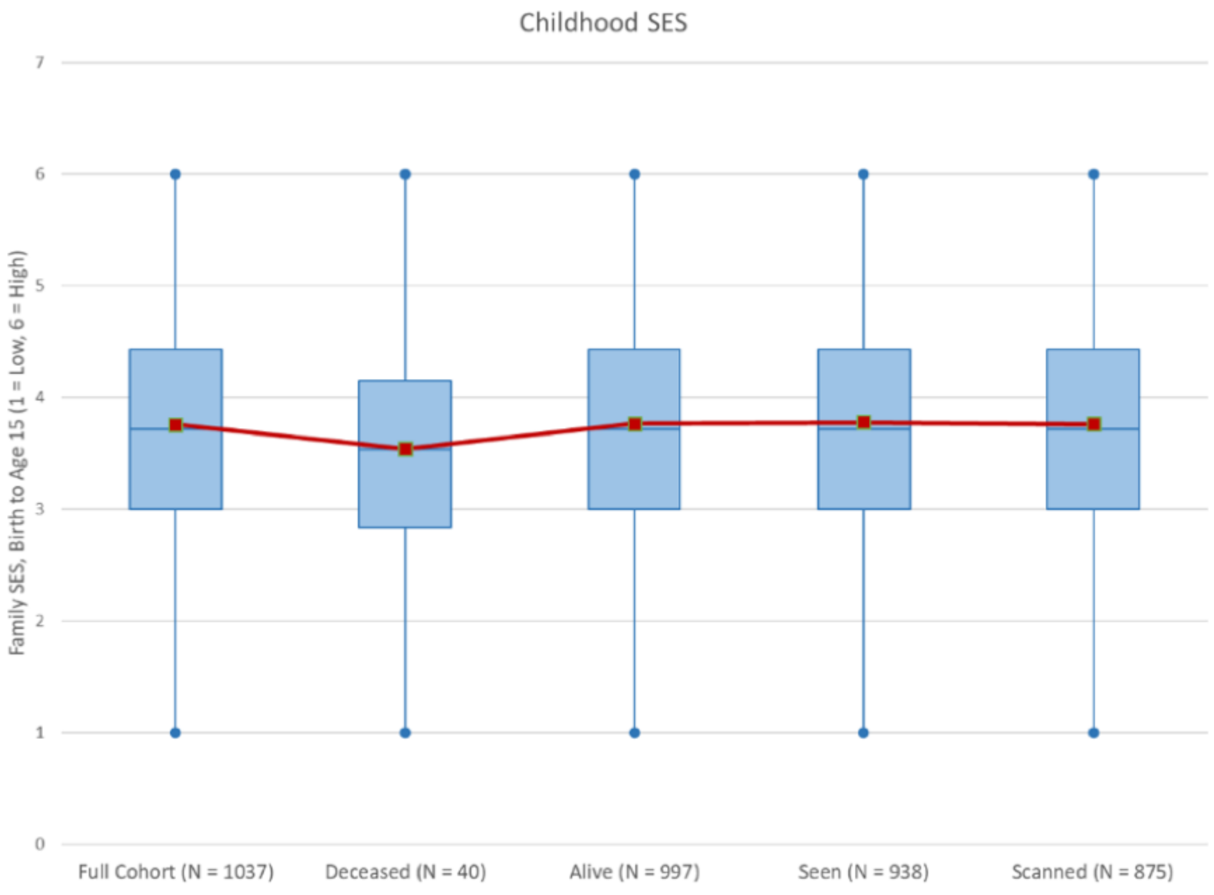

**Supplemental Figure S3. Testing biomarker change for nonlinearity.** For each biomarker, the graphs show raw age-26-standardized z-scores (blue lines) at each assessment age, estimated linear-only model (red lines), and estimated linear + quadratic model (purple lines). **Panel A** shows biomarkers for which fit statistics indicated that the linear model provided a better fit than the quadratic model. **Panel B** shows biomarkers for which fit statistics (-2 residual LL, AICC, BIC) indicated that the quadratic model provided a better fit than the linear model. However, for these seven biomarkers, the linear slope estimates extracted from the two models were highly correlated (Waist-hip ratio: 0.99, $VO_2$Max: 1.00, $FEV_1$/FVC: 0.99, $FEV_1$: 0.99, Apolipoprotein B100/A1 ratio: 0.99; BUN: 0.99; Gum health: 0.99), leading us to conclude that we could reasonably use the linear slope estimates from the models including linear fixed effects only. **Panel C** shows biomarkers that were only measured at only three time-points and, thus, could only be fit with a linear model.

**Panel A: Biomarkers for which linear model fit better than quadratic model.**

**BMI**

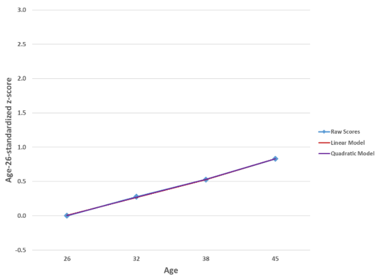


**HbA1c**

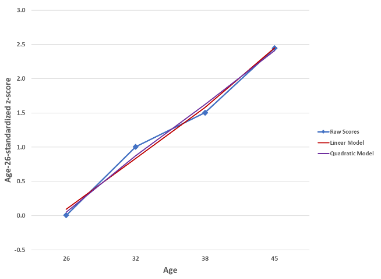


**Mean arterial pressure**

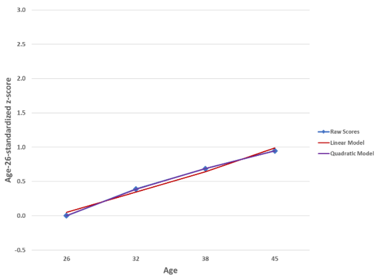


**Total cholesterol**

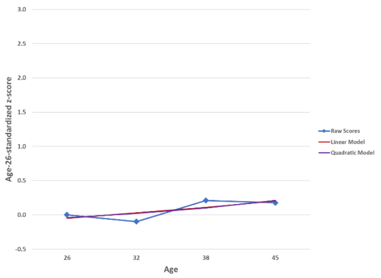


**Triglycerides**

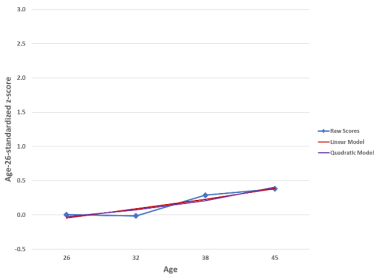


**HDL cholesterol (reversed)**

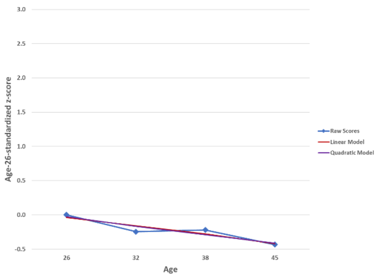


**Lipoprotein(a)**

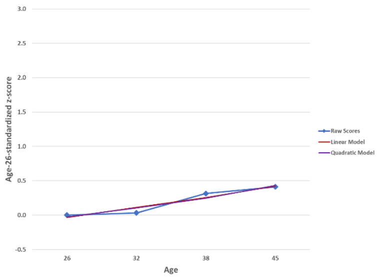


**White blood cell count**

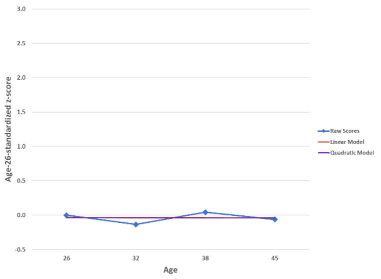


**Dental caries experience**

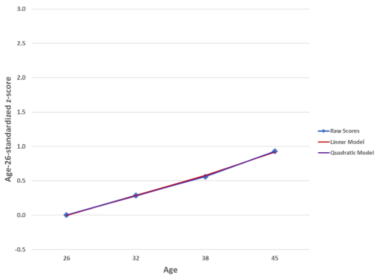

**Panel B: Biomarkers for which quadratic model fit better than linear model.**

### Waist-hip ratio

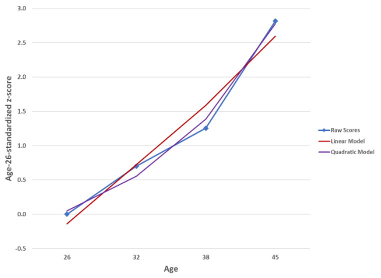


### $VO_2$Max (reversed)

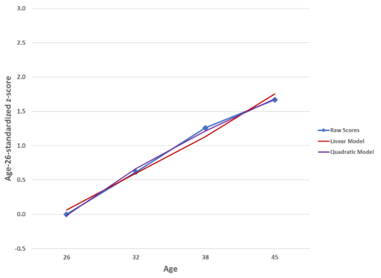


### $FEV_1$/FVC (reversed)

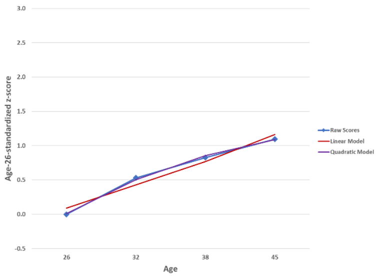


### $FEV_1$ (reversed)

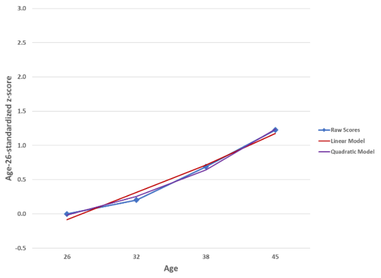


### ApoB100/ApoA1

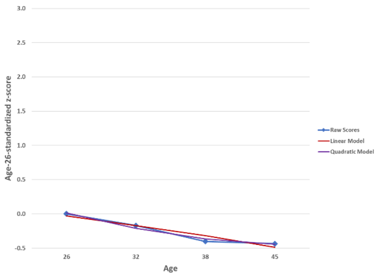


### Blood Urea Nitrogen

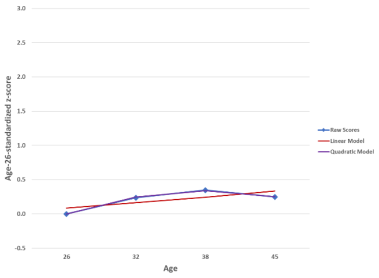


### Mean periodontal AL

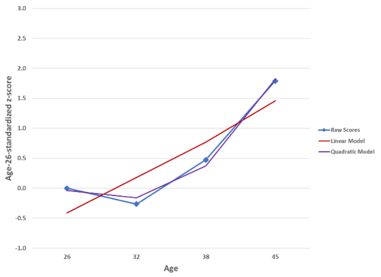

**Panel C: Biomarkers measured at 3 time points and not tested for non-linearity.**

**Leptin**

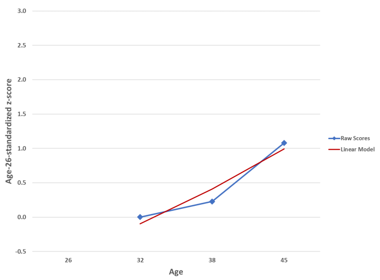


**hs-CRP**

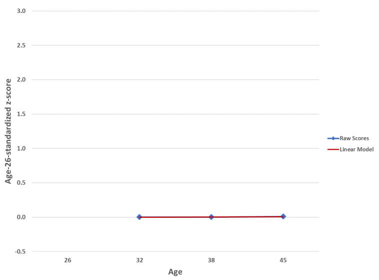


**eGFR (reversed)**

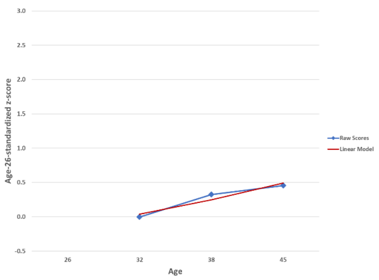

**Supplemental Table S1. Measurement of biomarkers used to calculate Pace of Aging measures.** Measures were taken in counterbalanced order across Study members with the exception of blood, which was drawn at the same time of day for all Study members at all four ages. Women who were pregnant at the time of a given assessment were excluded from that wave of data collection. In our previous 2015 article, we reported the pace of aging with 3 waves from 26 to 38 with 18 biomarkers. Here we add a 4th measurement wave of data completed in 2019, at age 45, with 19 biomarkers. We added measures of leptin and carries, both of which are now available at multiple waves allowing growth curve modeling, and we dropped telomere length because of an emerging and yet-unresolved field-wide debate about its measurement.

| | |
|---|---|
| ***Body mass index*** | Height was measured to the nearest millimeter using a portable stadiometer (Harpenden; Holtain, Ltd.). Weight was measured to the nearest 0.1 kg using calibrated scales. Individuals were weighed in light clothing. Body mass index (BMI) was calculated. |
| ***Waist-hip ratio*** | Waist girth was the perimeter at the level of the noticeable waist narrowing located between the costal border and the iliac crest. Hip girth was taken as the perimeter at the level of the greatest protuberance and at about the symphysion pubic level anteriorly. Measurements were repeated and the average used to calculate waist-hip ratio. |
| ***Glycated hemoglobin (HbA1C)*** | Whole blood glycated hemoglobin concentration (expressed as a percentage of total hemoglobin) was measured by ion exchange high performance liquid chromatography (Variant II: BioRad, Hercultes, Calif.), a method certified by the US National Glycohemoglobin Standardization Program (http://www.ngsp.org/). |
| ***Leptin*** | Serum leptin (µg/L) was measured using Human Leptin RIA kit (Cat# HL-81K, Linco Research, Missouri, USA) (Ages 32 & 38) and the Quantikine ELISA Human Leptin Immunoassay (Cat# SLP00, R&D Systems Inc, Minneapolis, MN) (Age 45) according to the manufacturer's instructions. |
| ***Blood pressure (mean arterial pressure)*** | Systolic and diastolic blood pressure were assessed according to standard protocols with a Hawksley random-zero sphygmomanometer with a constant deflation valve. Mean arterial pressure (MAP) was calculated using the formula Diastolic Pressure+1/3(Systolic Pressure - Diastolic Pressure). |
| ***Cardiorespiratory fitness ($VO_2$Max)*** | Cardiorespiratory fitness was assessed by measuring heart rate in response to a submaximal exercise test on a friction-braked cycle ergometer. Dependent on the extent to which heart rate increased during a 2-min 50 W warm-up, the workload was adjusted to elicit a steady heart-rate in the range 130–170 beats per minute. After a further 6-min constant power output stage, the maximum heart rate was recorded and used to calculate predicted maximum oxygen uptake adjusted for body weight in milliliters per minute per kilogram ($VO_2$max) according to standard protocols. |

| | |
|---|---|
| ***Lung function ($FEV_1$ and $FEV_1$/FVC)*** | We calculated post-albuterol forced expiratory volume in one second ($FEV_1$) and the ratio of $FEV_1$ to forced vital capacity (FVC; $FEV_1$/FVC) using measurements from spirometry conducted with a Sensormedics body plethysmograph (Sensormedics Corporation, Yorba Linda, CA, USA). |
| ***Total cholesterol, triglycerides, and high-density lipoprotein (HDL) cholesterol*** | Serum non-fasting total cholesterol, triglycerides, and high-density lipoprotein (HDL) cholesterol levels (mmol/L) were measured by colorimetric assay on a Hitachi 917 analyzer (ages 26-32), a Modular P analyzer (age 38), and a Cobas c702 analyzer (age 45). |
| ***Lipoprotein(a)*** | Serum lipoprotein(a) (nmol/L) was measured by a particle-enhanced immunoturbidimetric assay on a Hitachi 917 analyzer (ages 26-32), a Modular P analyzer (age 38), and a Cobas c502 analyzer (age 45). |
| ***Apolipoprotein B100/A1 ratio*** | Serum apolipoprotein A1 and apolipoprotein B100 (g/L) were measured by immunoturbidimetric assay on a Hitachi 917 analyzer (ages 26-32), a Modular P analyzer (age 38), and a Cobas c502 (age 45), and the ratio between the two was calculated. |
| ***eGFR (estimated glomerular filtration rate)*** | Serum creatinine (mmol/L) was measured by kinetic colorimetric assay on a Hitachi 917 analyzer (age 32), Modular P analyzer (age 38), and Cobas c702 (age 45) (Roche Diagnostics, Mannheim, Germany). Cystatin C was measured in Serum using an immunoturbidimetric assay implemented on the Cobas c 502 platform. The assay reagents kit used was CYSC2 Tina-quant Cystatin C Gen.2, Cat# 06600239 190, ACN 8109, Roche Diagnostics GmbH, D-68305 Mannheim, Germany. The calibrator kit was C.f.a.s. Cystatin C, Cat# 04975901 191, Roche Diagnostics GmbH, D-68305 Mannheim, Germany. Standardised against ERM-DA471/IFCC reference material. The lower limit of detection was 0.40mg/L. eGFR was estimated utilizing the CKD-Epi formula calculated from serum creatinine and serum cystatin C. |
| ***Blood urea nitrogen (BUN)*** | Blood urea nitrogen (mmol/L) was measured by kinetic UV assay at ages 26 (Hitachi 917 analyzer) and 45 (Cobas c702 analyzer), and by kinetic colorimetric assay at ages 32 (Hitachi 917 analyzer) and 38 (Modular P analyzer). |
| ***High sensitivity C-reactive protein (hs-CRP)*** | Serum C-reactive protein (mg/L) was measured by high sensitivity immunoturbidimetric assay on a Hitachi 917 analyzer (age 32), a Modular P analyzer (age 38), and a Cobas c702 (age 45). Values were log-transformed for analysis. |
| ***White blood cell count*** | Whole blood white blood cell counts ($x10^9$/L) were measured by flow cytometry with a Coulter STKS (Coulter Corporation, Miami, FL) (age 26), a Sysmex XE2100 (Sysmex Corporation, Japan) (age 32), and a Sysmex XE5000 (Sysmex Corporation, Japan) (ages 38 and 45). Counts were log-transformed for analysis. |

| | |
|---|---|
| ***Mean periodontal attachment loss (AL)*** | Calibrated dentists used a PCP-2 periodontal probe (Hu-Friedy; Chicago) to measure gingival recession (the distance from the cementoenamel junction to the gingival margin) and probing depth (the distance from the probe tip to the gingival margin) in millimeters at three sites (mesiobuccal, buccal, and distolingual) per tooth, excluding third molar teeth. Periodontal attachment loss for each site was computed by summing gingival recession and probing depth. and then averaged across all periodontally examined teeth. Periodontal examinations were conducted with half-mouth examinations at age 26 and full-mouth examinations at ages 32, 38, and 45 years. |
| ***Caries-affected tooth surfaces*** | Calibrated dentists examined the teeth for caries and restorations following the World Health Organization Oral Health Surveys methodology. Four surfaces were considered for anterior teeth (canines and incisors): buccal, lingual, distal, and mesial; a fifth surface, occlusal, was considered for premolar and molar teeth. Tooth surfaces were classified as having untreated caries (DS) if a cavitated carious lesion was present, as filled (FS) if a dental restoration was present (including crowns), and missing due to caries (MS) if the participant indicated that a given tooth had been removed due to decay or failed dental restorative work. DS, MS, and FS counts were summed to obtain a DMFS score (ranging from 0 to 148 surfaces). Caries experience was expressed as the % of tooth surfaces that had been caries-affected, excluding surfaces of teeth that were unerupted, lost due to trauma, extracted for reasons other than caries (impaction, orthodontic treatment, or periodontal disease), or could not be visualized by the examiner. |

**Supplemental Table S2. Pairwise correlations among Study-member specific slopes for 19 biomarkers.**

| | BMI | Waist-hip ratio | HbA1c | Leptin | Mean arterial pressure | $VO_2$Max (rev) | $FEV_1$/FVC (rev) | $FEV_1$ | Total cholesterol | Triglycerides | HDL Cholesterol (rev) | Lipoprotein(a) | ApoB100/ApoA1 | eGFR (rev) | Blood urea nitrogen | hs-CRP | White blood cell count | Mean periodontal AL | Dental caries experience |
|---|---|---|---|---|---|---|---|---|---|---|---|---|---|---|---|---|---|---|---|
| Mean slope | .04 | .14 | .12 | .08 | .05 | .09 | .06 | .07 | .01 | .02 | -.02 | .02 | -.02 | .03 | .01 | .00 | .00 | .10 | .05 |
| BMI | -- | | | | | | | | | | | | | | | | | | |
| Waist-hip ratio | .4 | -- | | | | | | | | | | | | | | | | | |
| HbA1c | .2 | .2 | -- | | | | | | | | | | | | | | | | |
| Leptin | .7 | .2 | .1 | -- | | | | | | | | | | | | | | | |
| Mean arterial pressure | .4 | .2 | .1 | .3 | -- | | | | | | | | | | | | | | |
| $VO_2$Max (rev) | .4 | .4 | .3 | .0 | .2 | -- | | | | | | | | | | | | | |
| $FEV_1$/FVC (rev) | -.2 | -.2 | -.1 | -.1 | -.1 | -.2 | -- | | | | | | | | | | | | |
| $FEV_1$ (rev) | .1 | .2 | .1 | -.1 | .0 | .2 | .3 | -- | | | | | | | | | | | |
| Total cholesterol | .2 | .2 | .1 | -.1 | .1 | .6 | -.2 | .1 | -- | | | | | | | | | | |
| Triglycerides | .4 | .4 | .2 | .1 | .2 | .3 | -.1 | .2 | .3 | -- | | | | | | | | | |
| HDL cholesterol (rev) | .4 | .3 | .1 | .2 | .1 | .2 | .0 | .1 | .0 | .5 | -- | | | | | | | | |
| Lipoprotein(a) | .1 | .0 | .0 | .0 | .0 | .0 | -.1 | .0 | .1 | .0 | -.1 | -- | | | | | | | |
| ApoB100/ApoA1 | .3 | .4 | .3 | .0 | .1 | .7 | -.2 | .3 | .7 | .4 | .4 | .1 | -- | | | | | | |
| eGFR (rev) | .0 | -.1 | .0 | .1 | .0 | -.2 | .0 | .0 | -.2 | -.1 | .0 | .0 | -.1 | -- | | | | | |
| Blood urea nitrogen | -.1 | -.1 | .0 | .1 | .0 | -.2 | .0 | -.1 | -.1 | .0 | .0 | .0 | -.2 | .4 | -- | | | | |
| hs-CRP | .3 | .2 | .1 | .2 | .2 | .2 | .0 | .1 | .1 | .1 | .2 | .0 | .2 | .0 | .0 | -- | | | |
| White blood cell count | .2 | .1 | .1 | .2 | .2 | .1 | .1 | .1 | .1 | .2 | .2 | .0 | .1 | .1 | .0 | .3 | -- | | |
| Mean periodontal AL | .0 | .1 | .1 | .0 | .0 | .1 | .2 | .2 | .0 | .1 | .1 | .0 | .1 | .0 | -.1 | .1 | .1 | -- | |
| Dental caries experience | .0 | .1 | .1 | .0 | .0 | .1 | .2 | .2 | .0 | .1 | .1 | .0 | .1 | .0 | -.1 | .1 | .1 | .5 | -- |

**Supplemental Table S3. Parcel-wise cortical thickness results.** Sex-adjusted associations between the Pace of Aging and cortical thickness for each of the cortical parcels in the Glasser atlas. The table reports standardized betas, p-values, and p-values that have been adjusted for 360 comparisons using the false discovery rate.

| Parcel label | beta | P | Adjusted P |
|---|---|---|---|
| R_V1_ROI | 0.015 | 0.652 | 0.718 |
| R_MST_ROI | -0.063 | 0.066 | 0.124 |
| R_V6_ROI | -0.088 | 0.010 | 0.030 |
| R_V2_ROI | -0.005 | 0.884 | 0.910 |
| R_V3_ROI | -0.051 | 0.138 | 0.213 |
| R_V4_ROI | -0.069 | 0.043 | 0.089 |
| R_V8_ROI | -0.070 | 0.041 | 0.089 |
| R_4_ROI | -0.105 | 0.002 | 0.008 |
| R_3b_ROI | 0.056 | 0.101 | 0.168 |
| R_FEF_ROI | -0.062 | 0.069 | 0.126 |
| R_PEF_ROI | -0.033 | 0.333 | 0.431 |
| R_55b_ROI | -0.023 | 0.503 | 0.588 |
| R_V3A_ROI | -0.058 | 0.090 | 0.154 |
| R_RSC_ROI | -0.003 | 0.931 | 0.952 |
| R_POS2_ROI | -0.104 | 0.002 | 0.009 |
| R_V7_ROI | -0.074 | 0.029 | 0.070 |
| R_IPS1_ROI | -0.065 | 0.054 | 0.104 |
| R_FFC_ROI | -0.056 | 0.100 | 0.168 |
| R_V3B_ROI | -0.072 | 0.032 | 0.074 |
| R_LO1_ROI | 0.007 | 0.835 | 0.874 |
| R_LO2_ROI | 0.002 | 0.956 | 0.970 |
| R_PIT_ROI | -0.041 | 0.227 | 0.329 |
| R_MT_ROI | -0.080 | 0.019 | 0.050 |
| R_A1_ROI | -0.069 | 0.043 | 0.089 |
| R_PSL_ROI | -0.024 | 0.486 | 0.573 |
| R_SFL_ROI | -0.019 | 0.575 | 0.646 |
| R_PCV_ROI | -0.025 | 0.462 | 0.555 |
| R_STV_ROI | -0.069 | 0.042 | 0.089 |
| R_7Pm_ROI | -0.041 | 0.234 | 0.337 |
| R_7m_ROI | -0.124 | 0.000 | 0.002 |
| R_POS1_ROI | -0.150 | 0.000 | 0.000 |
| R_23d_ROI | -0.085 | 0.011 | 0.032 |
| R_v23ab_ROI | -0.076 | 0.022 | 0.056 |
| R_d23ab_ROI | -0.089 | 0.009 | 0.028 |
| R_31pv_ROI | -0.066 | 0.054 | 0.104 |
| R_5m_ROI | 0.005 | 0.885 | 0.910 |
| R_5mv_ROI | -0.002 | 0.954 | 0.970 |
| R_23c_ROI | -0.098 | 0.003 | 0.013 |
| R_5L_ROI | 0.070 | 0.039 | 0.085 |
| R_24dd_ROI | -0.013 | 0.703 | 0.764 |
| R_24dv_ROI | -0.024 | 0.488 | 0.573 |
| R_7AL_ROI | 0.024 | 0.488 | 0.573 |
| R_SCEF_ROI | 0.032 | 0.343 | 0.437 |
| R_6ma_ROI | -0.036 | 0.290 | 0.389 |

| | | | |
|---|---|---|---|
| R_7Am_ROI | 0.035 | 0.304 | 0.399 |
| R_7PL_ROI | 0.000 | 0.994 | 0.998 |
| R_7PC_ROI | 0.040 | 0.239 | 0.340 |
| R_LIPv_ROI | 0.001 | 0.982 | 0.990 |
| R_VIP_ROI | 0.019 | 0.583 | 0.652 |
| R_MIP_ROI | -0.002 | 0.956 | 0.970 |
| R_1_ROI | -0.001 | 0.969 | 0.980 |
| R_2_ROI | -0.052 | 0.124 | 0.199 |
| R_3a_ROI | -0.052 | 0.131 | 0.206 |
| R_6d_ROI | -0.022 | 0.523 | 0.606 |
| R_6mp_ROI | -0.042 | 0.223 | 0.327 |
| R_6v_ROI | -0.018 | 0.596 | 0.665 |
| R_p24pr_ROI | -0.086 | 0.011 | 0.033 |
| R_33pr_ROI | 0.013 | 0.697 | 0.760 |
| R_a24pr_ROI | -0.059 | 0.082 | 0.144 |
| R_p32pr_ROI | -0.040 | 0.242 | 0.341 |
| R_a24_ROI | -0.051 | 0.136 | 0.210 |
| R_d32_ROI | -0.063 | 0.064 | 0.120 |
| R_8BM_ROI | 0.017 | 0.624 | 0.694 |
| R_p32_ROI | -0.033 | 0.337 | 0.432 |
| R_10r_ROI | -0.025 | 0.469 | 0.561 |
| R_47m_ROI | -0.110 | 0.001 | 0.006 |
| R_8Av_ROI | -0.049 | 0.154 | 0.235 |
| R_8Ad_ROI | -0.020 | 0.550 | 0.626 |
| R_9m_ROI | -0.019 | 0.576 | 0.646 |
| R_8BL_ROI | 0.007 | 0.833 | 0.874 |
| R_9p_ROI | 0.038 | 0.263 | 0.362 |
| R_10d_ROI | 0.021 | 0.531 | 0.612 |
| R_8C_ROI | -0.005 | 0.894 | 0.917 |
| R_44_ROI | -0.058 | 0.086 | 0.150 |
| R_45_ROI | -0.074 | 0.030 | 0.070 |
| R_47l_ROI | -0.098 | 0.004 | 0.014 |
| R_a47r_ROI | -0.052 | 0.130 | 0.205 |
| R_6r_ROI | -0.068 | 0.042 | 0.089 |
| R_IFJa_ROI | -0.037 | 0.281 | 0.379 |
| R_IFJp_ROI | -0.062 | 0.070 | 0.127 |
| R_IFSp_ROI | -0.021 | 0.545 | 0.622 |
| R_IFSa_ROI | -0.029 | 0.386 | 0.474 |
| R_p9-46v_ROI | -0.059 | 0.081 | 0.144 |
| R_46_ROI | -0.055 | 0.109 | 0.179 |
| R_a9-46v_ROI | -0.021 | 0.534 | 0.613 |
| R_9-46d_ROI | -0.035 | 0.298 | 0.395 |
| R_9a_ROI | 0.020 | 0.567 | 0.642 |
| R_10v_ROI | -0.105 | 0.002 | 0.008 |
| R_a10p_ROI | -0.011 | 0.743 | 0.799 |
| R_10pp_ROI | -0.083 | 0.015 | 0.041 |
| R_11l_ROI | -0.078 | 0.022 | 0.055 |
| R_13l_ROI | -0.211 | 0.000 | 0.000 |
| R_OFC_ROI | -0.058 | 0.089 | 0.153 |
| R_47s_ROI | -0.203 | 0.000 | 0.000 |

| | | | |
|---|---|---|---|
| R_LIPd_ROI | 0.069 | 0.045 | 0.092 |
| R_6a_ROI | -0.033 | 0.337 | 0.432 |
| R_i6-8_ROI | -0.043 | 0.208 | 0.307 |
| R_s6-8_ROI | 0.006 | 0.870 | 0.900 |
| R_43_ROI | -0.047 | 0.164 | 0.248 |
| R_OP4_ROI | -0.062 | 0.068 | 0.125 |
| R_OP1_ROI | -0.100 | 0.004 | 0.013 |
| R_OP2-3_ROI | -0.063 | 0.059 | 0.112 |
| R_52_ROI | -0.037 | 0.267 | 0.366 |
| R_RI_ROI | -0.070 | 0.039 | 0.086 |
| R_PFcm_ROI | -0.099 | 0.004 | 0.013 |
| R_PoI2_ROI | -0.124 | 0.000 | 0.001 |
| R_TA2_ROI | -0.142 | 0.000 | 0.000 |
| R_FOP4_ROI | -0.121 | 0.000 | 0.002 |
| R_MI_ROI | -0.092 | 0.007 | 0.023 |
| R_Pir_ROI | -0.059 | 0.084 | 0.147 |
| R_AVI_ROI | -0.133 | 0.000 | 0.001 |
| R_AAIC_ROI | -0.154 | 0.000 | 0.000 |
| R_FOP1_ROI | -0.102 | 0.002 | 0.009 |
| R_FOP3_ROI | -0.122 | 0.000 | 0.002 |
| R_FOP2_ROI | -0.040 | 0.241 | 0.340 |
| R_PFt_ROI | -0.042 | 0.223 | 0.327 |
| R_AIP_ROI | -0.038 | 0.271 | 0.371 |
| R_EC_ROI | -0.124 | 0.000 | 0.002 |
| R_PreS_ROI | 0.006 | 0.851 | 0.886 |
| R_H_ROI | -0.132 | 0.000 | 0.001 |
| R_ProS_ROI | -0.082 | 0.014 | 0.039 |
| R_PeEc_ROI | -0.218 | 0.000 | 0.000 |
| R_STGa_ROI | -0.118 | 0.001 | 0.003 |
| R_PBelt_ROI | -0.109 | 0.001 | 0.006 |
| R_A5_ROI | -0.144 | 0.000 | 0.000 |
| R_PHA1_ROI | -0.088 | 0.010 | 0.029 |
| R_PHA3_ROI | -0.127 | 0.000 | 0.001 |
| R_STSda_ROI | -0.175 | 0.000 | 0.000 |
| R_STSdp_ROI | -0.093 | 0.006 | 0.019 |
| R_STSvp_ROI | -0.047 | 0.162 | 0.246 |
| R_TGd_ROI | -0.202 | 0.000 | 0.000 |
| R_TE1a_ROI | -0.061 | 0.068 | 0.125 |
| R_TE1p_ROI | 0.029 | 0.399 | 0.486 |
| R_TE2a_ROI | -0.098 | 0.004 | 0.013 |
| R_TF_ROI | -0.137 | 0.000 | 0.000 |
| R_TE2p_ROI | -0.087 | 0.011 | 0.032 |
| R_PHT_ROI | -0.026 | 0.440 | 0.530 |
| R_PH_ROI | -0.139 | 0.000 | 0.000 |
| R_TPOJ1_ROI | -0.032 | 0.352 | 0.446 |
| R_TPOJ2_ROI | -0.074 | 0.030 | 0.070 |
| R_TPOJ3_ROI | -0.058 | 0.084 | 0.147 |
| R_DVT_ROI | -0.101 | 0.003 | 0.012 |
| R_PGp_ROI | -0.047 | 0.167 | 0.250 |
| R_IP2_ROI | -0.054 | 0.117 | 0.189 |

| | | | |
|---|---|---|---|
| R_IP1_ROI | 0.011 | 0.758 | 0.813 |
| R_IP0_ROI | -0.039 | 0.251 | 0.349 |
| R_PFop_ROI | -0.027 | 0.423 | 0.515 |
| R_PF_ROI | -0.033 | 0.336 | 0.432 |
| R_PFm_ROI | -0.008 | 0.813 | 0.856 |
| R_PGi_ROI | -0.055 | 0.108 | 0.177 |
| R_PGs_ROI | -0.014 | 0.684 | 0.749 |
| R_V6A_ROI | -0.119 | 0.001 | 0.003 |
| R_VMV1_ROI | -0.070 | 0.039 | 0.086 |
| R_VMV3_ROI | -0.089 | 0.008 | 0.026 |
| R_PHA2_ROI | -0.173 | 0.000 | 0.000 |
| R_V4t_ROI | -0.032 | 0.354 | 0.447 |
| R_FST_ROI | -0.083 | 0.014 | 0.039 |
| R_V3CD_ROI | -0.035 | 0.310 | 0.406 |
| R_LO3_ROI | -0.016 | 0.641 | 0.710 |
| R_VMV2_ROI | -0.129 | 0.000 | 0.001 |
| R_31pd_ROI | -0.051 | 0.135 | 0.210 |
| R_31a_ROI | -0.041 | 0.227 | 0.329 |
| R_VVC_ROI | -0.095 | 0.005 | 0.016 |
| R_25_ROI | -0.064 | 0.053 | 0.104 |
| R_s32_ROI | -0.032 | 0.350 | 0.446 |
| R_pOFC_ROI | -0.185 | 0.000 | 0.000 |
| R_PoI1_ROI | -0.080 | 0.016 | 0.042 |
| R_Ig_ROI | -0.030 | 0.377 | 0.469 |
| R_FOP5_ROI | -0.085 | 0.013 | 0.036 |
| R_p10p_ROI | -0.037 | 0.279 | 0.379 |
| R_p47r_ROI | -0.049 | 0.154 | 0.235 |
| R_TGv_ROI | -0.199 | 0.000 | 0.000 |
| R_MBelt_ROI | -0.132 | 0.000 | 0.001 |
| R_LBelt_ROI | -0.110 | 0.001 | 0.006 |
| R_A4_ROI | -0.074 | 0.030 | 0.070 |
| R_STSva_ROI | -0.091 | 0.006 | 0.020 |
| R_TE1m_ROI | -0.027 | 0.428 | 0.519 |
| R_PI_ROI | -0.126 | 0.000 | 0.001 |
| R_a32pr_ROI | -0.081 | 0.018 | 0.046 |
| R_p24_ROI | -0.055 | 0.107 | 0.177 |
| L_V1_ROI | 0.024 | 0.485 | 0.573 |
| L_MST_ROI | -0.030 | 0.384 | 0.474 |
| L_V6_ROI | -0.061 | 0.073 | 0.131 |
| L_V2_ROI | -0.019 | 0.574 | 0.646 |
| L_V3_ROI | -0.055 | 0.108 | 0.177 |
| L_V4_ROI | -0.108 | 0.001 | 0.006 |
| L_V8_ROI | -0.031 | 0.364 | 0.459 |
| L_4_ROI | -0.143 | 0.000 | 0.000 |
| L_3b_ROI | 0.077 | 0.024 | 0.059 |
| L_FEF_ROI | -0.131 | 0.000 | 0.001 |
| L_PEF_ROI | -0.109 | 0.001 | 0.006 |
| L_55b_ROI | -0.133 | 0.000 | 0.001 |
| L_V3A_ROI | -0.079 | 0.020 | 0.051 |
| L_RSC_ROI | -0.089 | 0.009 | 0.028 |

| | | | |
|---|---|---|---|
| L_POS2_ROI | -0.066 | 0.051 | 0.102 |
| L_V7_ROI | -0.045 | 0.188 | 0.279 |
| L_IPS1_ROI | -0.078 | 0.022 | 0.056 |
| L_FFC_ROI | -0.068 | 0.045 | 0.092 |
| L_V3B_ROI | -0.111 | 0.001 | 0.005 |
| L_LO1_ROI | -0.084 | 0.014 | 0.039 |
| L_LO2_ROI | -0.055 | 0.111 | 0.180 |
| L_PIT_ROI | -0.036 | 0.296 | 0.395 |
| L_MT_ROI | -0.022 | 0.511 | 0.596 |
| L_A1_ROI | -0.089 | 0.010 | 0.029 |
| L_PSL_ROI | -0.100 | 0.003 | 0.013 |
| L_SFL_ROI | 0.021 | 0.535 | 0.613 |
| L_PCV_ROI | -0.035 | 0.299 | 0.395 |
| L_STV_ROI | 0.009 | 0.801 | 0.848 |
| L_7Pm_ROI | 0.026 | 0.440 | 0.530 |
| L_7m_ROI | -0.076 | 0.026 | 0.063 |
| L_POS1_ROI | -0.134 | 0.000 | 0.001 |
| L_23d_ROI | -0.036 | 0.280 | 0.379 |
| L_v23ab_ROI | -0.133 | 0.000 | 0.001 |
| L_d23ab_ROI | -0.126 | 0.000 | 0.001 |
| L_31pv_ROI | -0.117 | 0.001 | 0.003 |
| L_5m_ROI | -0.012 | 0.722 | 0.781 |
| L_5mv_ROI | 0.031 | 0.371 | 0.463 |
| L_23c_ROI | -0.065 | 0.056 | 0.107 |
| L_5L_ROI | 0.067 | 0.050 | 0.100 |
| L_24dd_ROI | -0.039 | 0.249 | 0.347 |
| L_24dv_ROI | -0.068 | 0.047 | 0.096 |
| L_7AL_ROI | 0.066 | 0.053 | 0.103 |
| L_SCEF_ROI | -0.010 | 0.768 | 0.818 |
| L_6ma_ROI | 0.000 | 0.996 | 0.998 |
| L_7Am_ROI | 0.053 | 0.122 | 0.197 |
| L_7PL_ROI | -0.068 | 0.048 | 0.097 |
| L_7PC_ROI | -0.006 | 0.865 | 0.897 |
| L_LIPv_ROI | 0.010 | 0.770 | 0.818 |
| L_VIP_ROI | 0.010 | 0.764 | 0.816 |
| L_MIP_ROI | -0.041 | 0.227 | 0.329 |
| L_1_ROI | -0.073 | 0.032 | 0.074 |
| L_2_ROI | -0.030 | 0.384 | 0.474 |
| L_3a_ROI | -0.031 | 0.371 | 0.463 |
| L_6d_ROI | -0.073 | 0.033 | 0.076 |
| L_6mp_ROI | -0.112 | 0.001 | 0.005 |
| L_6v_ROI | -0.029 | 0.389 | 0.477 |
| L_p24pr_ROI | -0.068 | 0.046 | 0.094 |
| L_33pr_ROI | 0.024 | 0.478 | 0.570 |
| L_a24pr_ROI | -0.058 | 0.091 | 0.155 |
| L_p32pr_ROI | -0.092 | 0.007 | 0.023 |
| L_a24_ROI | -0.092 | 0.007 | 0.023 |
| L_d32_ROI | -0.070 | 0.040 | 0.087 |
| L_8BM_ROI | 0.016 | 0.647 | 0.715 |
| L_p32_ROI | -0.057 | 0.094 | 0.159 |

| | | | |
|---|---|---|---|
| L_10r_ROI | -0.036 | 0.297 | 0.395 |
| L_47m_ROI | -0.133 | 0.000 | 0.001 |
| L_8Av_ROI | -0.103 | 0.003 | 0.010 |
| L_8Ad_ROI | -0.054 | 0.113 | 0.183 |
| L_9m_ROI | -0.034 | 0.322 | 0.419 |
| L_8BL_ROI | -0.007 | 0.840 | 0.876 |
| L_9p_ROI | -0.020 | 0.561 | 0.637 |
| L_10d_ROI | -0.014 | 0.677 | 0.743 |
| L_8C_ROI | -0.073 | 0.033 | 0.075 |
| L_44_ROI | -0.037 | 0.275 | 0.376 |
| L_45_ROI | -0.070 | 0.040 | 0.086 |
| L_47l_ROI | -0.145 | 0.000 | 0.000 |
| L_a47r_ROI | -0.064 | 0.062 | 0.117 |
| L_6r_ROI | -0.124 | 0.000 | 0.001 |
| L_IFJa_ROI | -0.077 | 0.024 | 0.058 |
| L_IFJp_ROI | -0.116 | 0.001 | 0.003 |
| L_IFSp_ROI | -0.085 | 0.012 | 0.034 |
| L_IFSa_ROI | -0.092 | 0.007 | 0.022 |
| L_p9-46v_ROI | -0.062 | 0.070 | 0.127 |
| L_46_ROI | -0.024 | 0.488 | 0.573 |
| L_a9-46v_ROI | -0.034 | 0.317 | 0.413 |
| L_9-46d_ROI | -0.082 | 0.016 | 0.044 |
| L_9a_ROI | -0.040 | 0.241 | 0.340 |
| L_10v_ROI | -0.201 | 0.000 | 0.000 |
| L_a10p_ROI | -0.012 | 0.733 | 0.790 |
| L_10pp_ROI | -0.061 | 0.073 | 0.131 |
| L_11l_ROI | -0.112 | 0.001 | 0.005 |
| L_13l_ROI | -0.253 | 0.000 | 0.000 |
| L_OFC_ROI | -0.109 | 0.001 | 0.006 |
| L_47s_ROI | -0.191 | 0.000 | 0.000 |
| L_LIPd_ROI | -0.040 | 0.245 | 0.343 |
| L_6a_ROI | -0.110 | 0.001 | 0.006 |
| L_i6-8_ROI | -0.067 | 0.051 | 0.101 |
| L_s6-8_ROI | -0.013 | 0.705 | 0.765 |
| L_43_ROI | -0.035 | 0.302 | 0.399 |
| L_OP4_ROI | -0.058 | 0.087 | 0.150 |
| L_OP1_ROI | -0.112 | 0.001 | 0.005 |
| L_OP2-3_ROI | -0.076 | 0.023 | 0.056 |
| L_52_ROI | -0.075 | 0.021 | 0.054 |
| L_RI_ROI | -0.072 | 0.034 | 0.076 |
| L_PFcm_ROI | -0.129 | 0.000 | 0.001 |
| L_PoI2_ROI | -0.148 | 0.000 | 0.000 |
| L_TA2_ROI | -0.092 | 0.006 | 0.020 |
| L_FOP4_ROI | -0.115 | 0.001 | 0.003 |
| L_MI_ROI | -0.107 | 0.002 | 0.007 |
| L_Pir_ROI | -0.143 | 0.000 | 0.000 |
| L_AVI_ROI | -0.175 | 0.000 | 0.000 |
| L_AAIC_ROI | -0.134 | 0.000 | 0.001 |
| L_FOP1_ROI | -0.137 | 0.000 | 0.000 |
| L_FOP3_ROI | -0.098 | 0.004 | 0.013 |

| | | | |
|---|---|---|---|
| L_FOP2_ROI | -0.089 | 0.008 | 0.025 |
| L_PFt_ROI | -0.072 | 0.034 | 0.076 |
| L_AIP_ROI | -0.069 | 0.042 | 0.089 |
| L_EC_ROI | -0.146 | 0.000 | 0.000 |
| L_PreS_ROI | -0.040 | 0.239 | 0.340 |
| L_H_ROI | -0.187 | 0.000 | 0.000 |
| L_ProS_ROI | -0.059 | 0.075 | 0.133 |
| L_PeEc_ROI | -0.218 | 0.000 | 0.000 |
| L_STGa_ROI | -0.087 | 0.010 | 0.030 |
| L_PBelt_ROI | -0.052 | 0.132 | 0.207 |
| L_A5_ROI | -0.100 | 0.003 | 0.012 |
| L_PHA1_ROI | -0.111 | 0.001 | 0.006 |
| L_PHA3_ROI | -0.134 | 0.000 | 0.001 |
| L_STSda_ROI | -0.127 | 0.000 | 0.001 |
| L_STSdp_ROI | -0.049 | 0.147 | 0.226 |
| L_STSvp_ROI | -0.071 | 0.034 | 0.076 |
| L_TGd_ROI | -0.226 | 0.000 | 0.000 |
| L_TE1a_ROI | -0.057 | 0.088 | 0.151 |
| L_TE1p_ROI | -0.052 | 0.129 | 0.205 |
| L_TE2a_ROI | -0.045 | 0.176 | 0.263 |
| L_TF_ROI | -0.179 | 0.000 | 0.000 |
| L_TE2p_ROI | -0.117 | 0.001 | 0.003 |
| L_PHT_ROI | -0.040 | 0.241 | 0.340 |
| L_PH_ROI | -0.106 | 0.002 | 0.008 |
| L_TPOJ1_ROI | -0.067 | 0.051 | 0.101 |
| L_TPOJ2_ROI | -0.082 | 0.016 | 0.044 |
| L_TPOJ3_ROI | -0.111 | 0.001 | 0.006 |
| L_DVT_ROI | -0.045 | 0.188 | 0.279 |
| L_PGp_ROI | -0.090 | 0.008 | 0.025 |
| L_IP2_ROI | -0.038 | 0.261 | 0.362 |
| L_IP1_ROI | -0.030 | 0.378 | 0.469 |
| L_IP0_ROI | -0.080 | 0.019 | 0.049 |
| L_PFop_ROI | -0.081 | 0.017 | 0.044 |
| L_PF_ROI | -0.066 | 0.052 | 0.103 |
| L_PFm_ROI | -0.074 | 0.031 | 0.071 |
| L_PGi_ROI | -0.092 | 0.007 | 0.023 |
| L_PGs_ROI | -0.049 | 0.155 | 0.236 |
| L_V6A_ROI | -0.052 | 0.129 | 0.205 |
| L_VMV1_ROI | -0.103 | 0.003 | 0.010 |
| L_VMV3_ROI | -0.117 | 0.001 | 0.003 |
| L_PHA2_ROI | -0.102 | 0.003 | 0.010 |
| L_V4t_ROI | -0.008 | 0.807 | 0.852 |
| L_FST_ROI | -0.022 | 0.513 | 0.596 |
| L_V3CD_ROI | -0.149 | 0.000 | 0.000 |
| L_LO3_ROI | -0.057 | 0.092 | 0.156 |
| L_VMV2_ROI | -0.109 | 0.001 | 0.006 |
| L_31pd_ROI | -0.064 | 0.057 | 0.109 |
| L_31a_ROI | -0.121 | 0.000 | 0.003 |
| L_VVC_ROI | -0.118 | 0.000 | 0.003 |
| L_25_ROI | -0.121 | 0.000 | 0.002 |

| | | | |
|---|---|---|---|
| L_s32_ROI | -0.063 | 0.067 | 0.124 |
| L_pOFC_ROI | -0.169 | 0.000 | 0.000 |
| L_PoI1_ROI | -0.142 | 0.000 | 0.000 |
| L_Ig_ROI | -0.107 | 0.002 | 0.007 |
| L_FOP5_ROI | -0.131 | 0.000 | 0.001 |
| L_p10p_ROI | -0.107 | 0.002 | 0.007 |
| L_p47r_ROI | -0.078 | 0.021 | 0.054 |
| L_TGv_ROI | -0.209 | 0.000 | 0.000 |
| L_MBelt_ROI | -0.104 | 0.002 | 0.010 |
| L_LBelt_ROI | -0.115 | 0.001 | 0.004 |
| L_A4_ROI | -0.077 | 0.024 | 0.059 |
| L_STSva_ROI | -0.076 | 0.024 | 0.058 |
| L_TE1m_ROI | 0.000 | 0.999 | 0.999 |
| L_PI_ROI | -0.120 | 0.000 | 0.001 |
| L_a32pr_ROI | -0.085 | 0.013 | 0.037 |
| L_p24_ROI | -0.120 | 0.000 | 0.003 |

**Supplemental Table S4. Parcel-wise surface area results.** Sex-adjusted associations between the Pace of Aging and surface area for each of the cortical parcels in the Glasser atlas. The table reports standardized betas, p-values, and p-values that have been adjusted for 360 comparisons using the false discovery rate.

| Parcel label | beta | P | Adjusted P |
|---|---|---|---|
| R_V1_ROI | -0.10 | 0.001 | 0.007 |
| R_MST_ROI | -0.06 | 0.038 | 0.109 |
| R_V6_ROI | -0.05 | 0.086 | 0.182 |
| R_V2_ROI | -0.10 | 0.001 | 0.011 |
| R_V3_ROI | -0.10 | 0.000 | 0.006 |
| R_V4_ROI | -0.10 | 0.000 | 0.006 |
| R_V8_ROI | -0.08 | 0.007 | 0.037 |
| R_4_ROI | -0.05 | 0.075 | 0.170 |
| R_3b_ROI | -0.05 | 0.086 | 0.182 |
| R_FEF_ROI | -0.07 | 0.025 | 0.086 |
| R_PEF_ROI | -0.03 | 0.429 | 0.529 |
| R_55b_ROI | -0.05 | 0.109 | 0.208 |
| R_V3A_ROI | -0.11 | 0.000 | 0.003 |
| R_RSC_ROI | 0.01 | 0.874 | 0.914 |
| R_POS2_ROI | -0.05 | 0.139 | 0.236 |
| R_V7_ROI | -0.12 | 0.000 | 0.004 |
| R_IPS1_ROI | -0.09 | 0.005 | 0.028 |
| R_FFC_ROI | -0.13 | 0.000 | 0.003 |
| R_V3B_ROI | -0.05 | 0.111 | 0.210 |
| R_LO1_ROI | -0.06 | 0.038 | 0.109 |
| R_LO2_ROI | -0.09 | 0.003 | 0.022 |
| R_PIT_ROI | -0.10 | 0.001 | 0.013 |
| R_MT_ROI | -0.05 | 0.102 | 0.201 |
| R_A1_ROI | -0.04 | 0.238 | 0.340 |
| R_PSL_ROI | -0.07 | 0.023 | 0.082 |
| R_SFL_ROI | -0.04 | 0.157 | 0.262 |
| R_PCV_ROI | -0.05 | 0.123 | 0.223 |
| R_STV_ROI | -0.07 | 0.022 | 0.081 |
| R_7Pm_ROI | -0.06 | 0.048 | 0.127 |
| R_7m_ROI | -0.08 | 0.011 | 0.048 |
| R_POS1_ROI | -0.03 | 0.275 | 0.374 |
| R_23d_ROI | -0.07 | 0.030 | 0.097 |
| R_v23ab_ROI | -0.06 | 0.083 | 0.182 |
| R_d23ab_ROI | -0.07 | 0.047 | 0.125 |

| | | | |
|---|---|---|---|
| R_31pv_ROI | 0.00 | 0.940 | 0.959 |
| R_5m_ROI | -0.04 | 0.264 | 0.366 |
| R_5mv_ROI | -0.03 | 0.413 | 0.515 |
| R_23c_ROI | -0.01 | 0.753 | 0.810 |
| R_5L_ROI | -0.04 | 0.195 | 0.311 |
| R_24dd_ROI | -0.03 | 0.274 | 0.374 |
| R_24dv_ROI | -0.02 | 0.448 | 0.543 |
| R_7AL_ROI | -0.02 | 0.574 | 0.651 |
| R_SCEF_ROI | -0.04 | 0.223 | 0.326 |
| R_6ma_ROI | -0.03 | 0.400 | 0.500 |
| R_7Am_ROI | -0.01 | 0.831 | 0.880 |
| R_7PL_ROI | -0.02 | 0.520 | 0.610 |
| R_7PC_ROI | -0.08 | 0.014 | 0.059 |
| R_LIPv_ROI | -0.03 | 0.346 | 0.450 |
| R_VIP_ROI | -0.05 | 0.106 | 0.203 |
| R_MIP_ROI | -0.01 | 0.863 | 0.905 |
| R_1_ROI | -0.07 | 0.032 | 0.100 |
| R_2_ROI | -0.04 | 0.190 | 0.309 |
| R_3a_ROI | -0.01 | 0.691 | 0.761 |
| R_6d_ROI | -0.09 | 0.003 | 0.025 |
| R_6mp_ROI | -0.03 | 0.304 | 0.406 |
| R_6v_ROI | -0.03 | 0.265 | 0.366 |
| R_p24pr_ROI | -0.05 | 0.121 | 0.222 |
| R_33pr_ROI | -0.05 | 0.131 | 0.231 |
| R_a24pr_ROI | -0.07 | 0.042 | 0.116 |
| R_p32pr_ROI | 0.00 | 0.941 | 0.959 |
| R_a24_ROI | -0.06 | 0.068 | 0.162 |
| R_d32_ROI | -0.05 | 0.119 | 0.221 |
| R_8BM_ROI | -0.03 | 0.348 | 0.450 |
| R_p32_ROI | -0.01 | 0.641 | 0.712 |
| R_10r_ROI | -0.03 | 0.385 | 0.483 |
| R_47m_ROI | -0.02 | 0.527 | 0.614 |
| R_8Av_ROI | -0.05 | 0.085 | 0.182 |
| R_8Ad_ROI | -0.04 | 0.194 | 0.310 |
| R_9m_ROI | -0.05 | 0.121 | 0.222 |
| R_8BL_ROI | -0.03 | 0.243 | 0.344 |
| R_9p_ROI | -0.04 | 0.237 | 0.340 |
| R_10d_ROI | -0.05 | 0.102 | 0.201 |
| R_8C_ROI | -0.02 | 0.452 | 0.545 |

| | | | |
|---|---|---|---|
| R_44_ROI | -0.08 | 0.007 | 0.037 |
| R_45_ROI | -0.07 | 0.023 | 0.083 |
| R_47l_ROI | -0.10 | 0.001 | 0.009 |
| R_a47r_ROI | -0.06 | 0.041 | 0.115 |
| R_6r_ROI | -0.03 | 0.340 | 0.443 |
| R_IFJa_ROI | -0.04 | 0.213 | 0.321 |
| R_IFJp_ROI | -0.05 | 0.143 | 0.243 |
| R_IFSp_ROI | 0.00 | 0.996 | 0.996 |
| R_IFSa_ROI | -0.08 | 0.012 | 0.051 |
| R_p9-46v_ROI | -0.08 | 0.010 | 0.046 |
| R_46_ROI | -0.06 | 0.079 | 0.177 |
| R_a9-46v_ROI | -0.02 | 0.437 | 0.535 |
| R_9-46d_ROI | -0.04 | 0.218 | 0.323 |
| R_9a_ROI | -0.05 | 0.122 | 0.222 |
| R_10v_ROI | -0.04 | 0.138 | 0.236 |
| R_a10p_ROI | -0.06 | 0.036 | 0.108 |
| R_10pp_ROI | -0.05 | 0.065 | 0.159 |
| R_11l_ROI | -0.07 | 0.024 | 0.084 |
| R_13l_ROI | -0.06 | 0.037 | 0.108 |
| R_OFC_ROI | -0.08 | 0.005 | 0.029 |
| R_47s_ROI | -0.07 | 0.019 | 0.074 |
| R_LIPd_ROI | -0.03 | 0.275 | 0.374 |
| R_6a_ROI | -0.06 | 0.068 | 0.162 |
| R_i6-8_ROI | -0.07 | 0.031 | 0.099 |
| R_s6-8_ROI | -0.07 | 0.031 | 0.099 |
| R_43_ROI | -0.05 | 0.137 | 0.236 |
| R_OP4_ROI | -0.01 | 0.732 | 0.796 |
| R_OP1_ROI | -0.05 | 0.091 | 0.189 |
| R_OP2-3_ROI | -0.02 | 0.434 | 0.534 |
| R_52_ROI | -0.04 | 0.243 | 0.344 |
| R_RI_ROI | -0.04 | 0.180 | 0.297 |
| R_PFcm_ROI | -0.01 | 0.727 | 0.793 |
| R_PoI2_ROI | -0.02 | 0.471 | 0.565 |
| R_TA2_ROI | -0.05 | 0.079 | 0.177 |
| R_FOP4_ROI | -0.07 | 0.020 | 0.074 |
| R_MI_ROI | -0.10 | 0.001 | 0.011 |
| R_Pir_ROI | 0.00 | 0.995 | 0.996 |
| R_AVI_ROI | -0.08 | 0.004 | 0.027 |
| R_AAIC_ROI | -0.04 | 0.186 | 0.306 |

| | | | |
|---|---|---|---|
| R_FOP1_ROI | -0.09 | 0.004 | 0.027 |
| R_FOP3_ROI | -0.05 | 0.098 | 0.199 |
| R_FOP2_ROI | -0.05 | 0.097 | 0.198 |
| R_PFt_ROI | 0.00 | 0.891 | 0.927 |
| R_AIP_ROI | -0.02 | 0.542 | 0.623 |
| R_EC_ROI | -0.09 | 0.005 | 0.029 |
| R_PreS_ROI | -0.08 | 0.011 | 0.048 |
| R_H_ROI | -0.05 | 0.072 | 0.169 |
| R_ProS_ROI | -0.02 | 0.506 | 0.598 |
| R_PeEc_ROI | -0.08 | 0.006 | 0.034 |
| R_STGa_ROI | -0.06 | 0.074 | 0.169 |
| R_PBelt_ROI | -0.06 | 0.070 | 0.165 |
| R_A5_ROI | -0.05 | 0.074 | 0.169 |
| R_PHA1_ROI | -0.04 | 0.231 | 0.334 |
| R_PHA3_ROI | -0.09 | 0.004 | 0.028 |
| R_STSda_ROI | -0.07 | 0.036 | 0.108 |
| R_STSdp_ROI | -0.08 | 0.007 | 0.035 |
| R_STSvp_ROI | -0.05 | 0.085 | 0.182 |
| R_TGd_ROI | -0.08 | 0.004 | 0.028 |
| R_TE1a_ROI | -0.06 | 0.036 | 0.108 |
| R_TE1p_ROI | -0.11 | 0.000 | 0.003 |
| R_TE2a_ROI | -0.08 | 0.009 | 0.043 |
| R_TF_ROI | -0.04 | 0.151 | 0.255 |
| R_TE2p_ROI | -0.04 | 0.167 | 0.277 |
| R_PHT_ROI | -0.09 | 0.004 | 0.028 |
| R_PH_ROI | -0.06 | 0.053 | 0.137 |
| R_TPOJ1_ROI | -0.09 | 0.005 | 0.029 |
| R_TPOJ2_ROI | -0.08 | 0.015 | 0.063 |
| R_TPOJ3_ROI | -0.01 | 0.745 | 0.802 |
| R_DVT_ROI | -0.05 | 0.093 | 0.193 |
| R_PGp_ROI | -0.11 | 0.000 | 0.005 |
| R_IP2_ROI | -0.04 | 0.193 | 0.310 |
| R_IP1_ROI | -0.05 | 0.101 | 0.201 |
| R_IP0_ROI | -0.03 | 0.352 | 0.451 |
| R_PFop_ROI | -0.04 | 0.228 | 0.330 |
| R_PF_ROI | -0.06 | 0.060 | 0.148 |
| R_PFm_ROI | -0.07 | 0.026 | 0.086 |
| R_PGi_ROI | -0.11 | 0.000 | 0.005 |
| R_PGs_ROI | -0.05 | 0.101 | 0.201 |

| | | | |
|---|---|---|---|
| R_V6A_ROI | -0.11 | 0.001 | 0.007 |
| R_VMV1_ROI | -0.05 | 0.132 | 0.233 |
| R_VMV3_ROI | -0.06 | 0.047 | 0.125 |
| R_PHA2_ROI | -0.02 | 0.536 | 0.618 |
| R_V4t_ROI | -0.06 | 0.044 | 0.120 |
| R_FST_ROI | -0.08 | 0.014 | 0.057 |
| R_V3CD_ROI | -0.11 | 0.000 | 0.006 |
| R_LO3_ROI | -0.12 | 0.000 | 0.004 |
| R_VMV2_ROI | -0.09 | 0.006 | 0.032 |
| R_31pd_ROI | 0.00 | 0.954 | 0.965 |
| R_31a_ROI | -0.03 | 0.336 | 0.440 |
| R_VVC_ROI | -0.07 | 0.026 | 0.086 |
| R_25_ROI | -0.06 | 0.042 | 0.116 |
| R_s32_ROI | -0.02 | 0.418 | 0.519 |
| R_pOFC_ROI | -0.11 | 0.000 | 0.005 |
| R_PoI1_ROI | -0.03 | 0.314 | 0.417 |
| R_Ig_ROI | -0.08 | 0.018 | 0.070 |
| R_FOP5_ROI | -0.04 | 0.198 | 0.314 |
| R_p10p_ROI | -0.03 | 0.372 | 0.469 |
| R_p47r_ROI | -0.03 | 0.289 | 0.390 |
| R_TGv_ROI | -0.08 | 0.005 | 0.029 |
| R_MBelt_ROI | -0.02 | 0.442 | 0.539 |
| R_LBelt_ROI | -0.10 | 0.003 | 0.022 |
| R_A4_ROI | -0.07 | 0.020 | 0.074 |
| R_STSva_ROI | -0.03 | 0.381 | 0.479 |
| R_TE1m_ROI | -0.08 | 0.004 | 0.027 |
| R_PI_ROI | 0.00 | 0.935 | 0.959 |
| R_a32pr_ROI | -0.04 | 0.235 | 0.339 |
| R_p24_ROI | 0.00 | 0.950 | 0.963 |
| L_V1_ROI | -0.11 | 0.000 | 0.003 |
| L_MST_ROI | -0.04 | 0.219 | 0.323 |
| L_V6_ROI | -0.08 | 0.014 | 0.057 |
| L_V2_ROI | -0.12 | 0.000 | 0.003 |
| L_V3_ROI | -0.11 | 0.000 | 0.003 |
| L_V4_ROI | -0.12 | 0.000 | 0.003 |
| L_V8_ROI | -0.09 | 0.005 | 0.030 |
| L_4_ROI | -0.04 | 0.212 | 0.321 |
| L_3b_ROI | -0.09 | 0.003 | 0.022 |
| L_FEF_ROI | -0.07 | 0.036 | 0.108 |

| | | | |
|---|---|---|---|
| L_PEF_ROI | -0.04 | 0.208 | 0.319 |
| L_55b_ROI | -0.06 | 0.066 | 0.159 |
| L_V3A_ROI | -0.12 | 0.000 | 0.003 |
| L_RSC_ROI | -0.01 | 0.722 | 0.790 |
| L_POS2_ROI | -0.06 | 0.057 | 0.144 |
| L_V7_ROI | -0.08 | 0.018 | 0.070 |
| L_IPS1_ROI | -0.06 | 0.074 | 0.169 |
| L_FFC_ROI | -0.08 | 0.012 | 0.050 |
| L_V3B_ROI | -0.03 | 0.301 | 0.403 |
| L_LO1_ROI | -0.06 | 0.061 | 0.150 |
| L_LO2_ROI | -0.12 | 0.000 | 0.003 |
| L_PIT_ROI | -0.11 | 0.000 | 0.003 |
| L_MT_ROI | -0.05 | 0.096 | 0.198 |
| L_A1_ROI | -0.05 | 0.135 | 0.234 |
| L_PSL_ROI | -0.05 | 0.115 | 0.215 |
| L_SFL_ROI | -0.04 | 0.248 | 0.349 |
| L_PCV_ROI | -0.05 | 0.105 | 0.203 |
| L_STV_ROI | -0.07 | 0.032 | 0.100 |
| L_7Pm_ROI | -0.04 | 0.261 | 0.366 |
| L_7m_ROI | -0.06 | 0.038 | 0.109 |
| L_POS1_ROI | -0.06 | 0.051 | 0.131 |
| L_23d_ROI | -0.04 | 0.200 | 0.314 |
| L_v23ab_ROI | -0.04 | 0.256 | 0.360 |
| L_d23ab_ROI | -0.05 | 0.127 | 0.227 |
| L_31pv_ROI | -0.05 | 0.125 | 0.223 |
| L_5m_ROI | -0.05 | 0.103 | 0.202 |
| L_5mv_ROI | -0.03 | 0.331 | 0.435 |
| L_23c_ROI | -0.05 | 0.134 | 0.234 |
| L_5L_ROI | -0.07 | 0.025 | 0.086 |
| L_24dd_ROI | 0.00 | 0.882 | 0.921 |
| L_24dv_ROI | -0.04 | 0.263 | 0.366 |
| L_7AL_ROI | -0.05 | 0.090 | 0.188 |
| L_SCEF_ROI | -0.02 | 0.453 | 0.545 |
| L_6ma_ROI | -0.01 | 0.822 | 0.873 |
| L_7Am_ROI | -0.04 | 0.208 | 0.319 |
| L_7PL_ROI | -0.05 | 0.086 | 0.182 |
| L_7PC_ROI | -0.08 | 0.011 | 0.050 |
| L_LIPv_ROI | -0.01 | 0.804 | 0.857 |
| L_VIP_ROI | -0.11 | 0.001 | 0.008 |

| | | | |
|---|---|---|---|
| L_MIP_ROI | -0.02 | 0.605 | 0.680 |
| L_1_ROI | -0.08 | 0.012 | 0.051 |
| L_2_ROI | -0.06 | 0.056 | 0.143 |
| L_3a_ROI | -0.05 | 0.085 | 0.182 |
| L_6d_ROI | -0.03 | 0.265 | 0.366 |
| L_6mp_ROI | -0.07 | 0.021 | 0.077 |
| L_6v_ROI | -0.03 | 0.298 | 0.400 |
| L_p24pr_ROI | -0.02 | 0.552 | 0.631 |
| L_33pr_ROI | -0.02 | 0.615 | 0.689 |
| L_a24pr_ROI | -0.06 | 0.086 | 0.182 |
| L_p32pr_ROI | 0.00 | 0.968 | 0.976 |
| L_a24_ROI | -0.05 | 0.110 | 0.208 |
| L_d32_ROI | -0.01 | 0.784 | 0.838 |
| L_8BM_ROI | -0.02 | 0.506 | 0.598 |
| L_p32_ROI | 0.02 | 0.531 | 0.616 |
| L_10r_ROI | -0.06 | 0.045 | 0.120 |
| L_47m_ROI | -0.08 | 0.008 | 0.042 |
| L_8Av_ROI | -0.02 | 0.491 | 0.585 |
| L_8Ad_ROI | 0.00 | 0.894 | 0.927 |
| L_9m_ROI | -0.06 | 0.032 | 0.100 |
| L_8BL_ROI | -0.03 | 0.322 | 0.426 |
| L_9p_ROI | -0.01 | 0.736 | 0.798 |
| L_10d_ROI | -0.04 | 0.221 | 0.324 |
| L_8C_ROI | -0.01 | 0.665 | 0.734 |
| L_44_ROI | -0.02 | 0.524 | 0.613 |
| L_45_ROI | -0.07 | 0.020 | 0.075 |
| L_47l_ROI | -0.09 | 0.004 | 0.025 |
| L_a47r_ROI | -0.08 | 0.009 | 0.043 |
| L_6r_ROI | -0.04 | 0.191 | 0.309 |
| L_IFJa_ROI | -0.06 | 0.073 | 0.169 |
| L_IFJp_ROI | 0.01 | 0.849 | 0.896 |
| L_IFSp_ROI | -0.04 | 0.214 | 0.321 |
| L_IFSa_ROI | -0.04 | 0.205 | 0.319 |
| L_p9-46v_ROI | -0.03 | 0.350 | 0.451 |
| L_46_ROI | -0.01 | 0.716 | 0.786 |
| L_a9-46v_ROI | -0.07 | 0.029 | 0.094 |
| L_9-46d_ROI | -0.02 | 0.444 | 0.539 |
| L_9a_ROI | -0.03 | 0.267 | 0.367 |
| L_10v_ROI | -0.09 | 0.002 | 0.020 |

| | | | |
|---|---|---|---|
| L_a10p_ROI | -0.08 | 0.013 | 0.057 |
| L_10pp_ROI | -0.05 | 0.108 | 0.207 |
| L_11l_ROI | -0.07 | 0.025 | 0.086 |
| L_13l_ROI | -0.08 | 0.007 | 0.037 |
| L_OFC_ROI | -0.09 | 0.001 | 0.013 |
| L_47s_ROI | -0.10 | 0.001 | 0.013 |
| L_LIPd_ROI | 0.01 | 0.743 | 0.802 |
| L_6a_ROI | 0.02 | 0.573 | 0.651 |
| L_i6-8_ROI | -0.02 | 0.515 | 0.606 |
| L_s6-8_ROI | 0.00 | 0.947 | 0.963 |
| L_43_ROI | -0.03 | 0.279 | 0.377 |
| L_OP4_ROI | -0.04 | 0.200 | 0.314 |
| L_OP1_ROI | -0.07 | 0.036 | 0.108 |
| L_OP2-3_ROI | 0.02 | 0.545 | 0.625 |
| L_52_ROI | -0.05 | 0.118 | 0.221 |
| L_RI_ROI | -0.03 | 0.371 | 0.469 |
| L_PFcm_ROI | 0.01 | 0.853 | 0.898 |
| L_PoI2_ROI | -0.06 | 0.059 | 0.146 |
| L_TA2_ROI | -0.07 | 0.010 | 0.047 |
| L_FOP4_ROI | -0.06 | 0.045 | 0.120 |
| L_MI_ROI | -0.11 | 0.000 | 0.006 |
| L_Pir_ROI | -0.02 | 0.589 | 0.664 |
| L_AVI_ROI | -0.06 | 0.050 | 0.131 |
| L_AAIC_ROI | -0.01 | 0.763 | 0.817 |
| L_FOP1_ROI | -0.05 | 0.121 | 0.222 |
| L_FOP3_ROI | -0.04 | 0.150 | 0.254 |
| L_FOP2_ROI | -0.04 | 0.201 | 0.314 |
| L_PFt_ROI | -0.03 | 0.364 | 0.464 |
| L_AIP_ROI | -0.09 | 0.007 | 0.035 |
| L_EC_ROI | -0.13 | 0.000 | 0.003 |
| L_PreS_ROI | -0.06 | 0.086 | 0.182 |
| L_H_ROI | -0.05 | 0.104 | 0.203 |
| L_ProS_ROI | -0.04 | 0.212 | 0.321 |
| L_PeEc_ROI | -0.09 | 0.005 | 0.029 |
| L_STGa_ROI | -0.07 | 0.024 | 0.084 |
| L_PBelt_ROI | 0.00 | 0.991 | 0.996 |
| L_A5_ROI | -0.05 | 0.124 | 0.223 |
| L_PHA1_ROI | 0.02 | 0.619 | 0.692 |
| L_PHA3_ROI | -0.08 | 0.016 | 0.064 |

| | | | |
|---|---|---|---|
| L_STSda_ROI | -0.04 | 0.219 | 0.323 |
| L_STSdp_ROI | -0.02 | 0.532 | 0.616 |
| L_STSvp_ROI | -0.09 | 0.003 | 0.025 |
| L_TGd_ROI | -0.11 | 0.000 | 0.003 |
| L_TE1a_ROI | -0.09 | 0.003 | 0.022 |
| L_TE1p_ROI | -0.10 | 0.001 | 0.010 |
| L_TE2a_ROI | -0.11 | 0.000 | 0.004 |
| L_TF_ROI | -0.07 | 0.025 | 0.085 |
| L_TE2p_ROI | -0.11 | 0.001 | 0.007 |
| L_PHT_ROI | -0.09 | 0.004 | 0.027 |
| L_PH_ROI | -0.13 | 0.000 | 0.003 |
| L_TPOJ1_ROI | -0.03 | 0.352 | 0.451 |
| L_TPOJ2_ROI | -0.08 | 0.020 | 0.074 |
| L_TPOJ3_ROI | -0.06 | 0.074 | 0.169 |
| L_DVT_ROI | -0.04 | 0.188 | 0.307 |
| L_PGp_ROI | -0.12 | 0.000 | 0.004 |
| L_IP2_ROI | -0.04 | 0.217 | 0.323 |
| L_IP1_ROI | -0.05 | 0.101 | 0.201 |
| L_IP0_ROI | -0.06 | 0.038 | 0.109 |
| L_PFop_ROI | -0.01 | 0.625 | 0.696 |
| L_PF_ROI | -0.05 | 0.137 | 0.236 |
| L_PFm_ROI | -0.06 | 0.033 | 0.101 |
| L_PGi_ROI | -0.04 | 0.208 | 0.319 |
| L_PGs_ROI | -0.12 | 0.000 | 0.003 |
| L_V6A_ROI | -0.10 | 0.002 | 0.014 |
| L_VMV1_ROI | -0.11 | 0.001 | 0.007 |
| L_VMV3_ROI | -0.02 | 0.507 | 0.598 |
| L_PHA2_ROI | 0.02 | 0.486 | 0.581 |
| L_V4t_ROI | -0.08 | 0.009 | 0.043 |
| L_FST_ROI | -0.05 | 0.098 | 0.199 |
| L_V3CD_ROI | -0.10 | 0.001 | 0.011 |
| L_LO3_ROI | -0.10 | 0.003 | 0.022 |
| L_VMV2_ROI | -0.03 | 0.366 | 0.466 |
| L_31pd_ROI | 0.00 | 0.904 | 0.930 |
| L_31a_ROI | -0.03 | 0.329 | 0.434 |
| L_VVC_ROI | -0.06 | 0.058 | 0.146 |
| L_25_ROI | -0.04 | 0.208 | 0.319 |
| L_s32_ROI | -0.04 | 0.226 | 0.329 |
| L_pOFC_ROI | -0.09 | 0.006 | 0.034 |

| | | | |
|---|---|---|---|
| L_PoI1_ROI | 0.00 | 0.897 | 0.928 |
| L_Ig_ROI | -0.05 | 0.155 | 0.259 |
| L_FOP5_ROI | -0.04 | 0.193 | 0.310 |
| L_p10p_ROI | -0.01 | 0.663 | 0.734 |
| L_p47r_ROI | -0.02 | 0.588 | 0.664 |
| L_TGv_ROI | -0.12 | 0.000 | 0.003 |
| L_MBelt_ROI | 0.00 | 0.902 | 0.930 |
| L_LBelt_ROI | -0.06 | 0.044 | 0.120 |
| L_A4_ROI | -0.06 | 0.058 | 0.146 |
| L_STSva_ROI | -0.04 | 0.211 | 0.321 |
| L_TE1m_ROI | -0.10 | 0.001 | 0.007 |
| L_PI_ROI | -0.05 | 0.128 | 0.227 |
| L_a32pr_ROI | -0.03 | 0.420 | 0.519 |
| L_p24_ROI | -0.06 | 0.080 | 0.178 |
